# Learning the natural history of human disease with generative transformers

**DOI:** 10.1101/2024.06.07.24308553

**Authors:** Artem Shmatko, Alexander Wolfgang Jung, Kumar Gaurav, Søren Brunak, Laust Mortensen, Ewan Birney, Tom Fitzgerald, Moritz Gerstung

## Abstract

Decision-making in healthcare relies on the ability to understand patients’ past and current health state to predict, and ultimately change, their future course. Artificial intelligence (AI) methods promise to aid this task by learning patterns of disease progression from large corpora of health records to predict detailed outcomes for an individual. However, the potential of AI has not yet been fully investigated at scale.

Here, we modify the GPT (generative pretrained transformer) architecture to model the temporal progression and competing nature of human diseases in a population scale cohort. We train this model, termed Delphi-2M, on data from 0.4 million participants of the UK Biobank and validate it using external data from 1.9 million Danish individuals with no change in parameters.

Delphi-2M predicts the rates of more than 1,000 different ICD-10 coded diseases and death, conditional on each individual’s past disease history, age, sex and baseline lifestyle information, and with accuracy comparable to existing single-disease models. Delphi-2M’s generative nature also enables sampling future health trajectories at any point within an individual’s life course with outcomes across the entire disease spectrum. Sampled health trajectories provide meaningful estimates of future disease burden for up to 20 years and enable training AI models which have never seen actual data.

Explainable AI methods provide insights into Delphi-2M’s predictions, revealing temporal clusters of co-morbidities within and across different disease chapters and their time-dependent consequences on the future health course. These analyses, however, also reveal that biases underlying the available training data, which in the case of the UK Biobank stem from distinct healthcare sources, are learned and highlighted.

In summary, GPT-based models appear well suited for predictive and generative health-related tasks, are applicable to population scale health data sets and provide insights into the temporal dependencies of past events that shape future health, impacting our ability to obtain an instantaneous view of personalised health state.

## Introduction

The progression of human disease across age is characterised by periods of health, episodes of acute illness and also chronic debilitation, often manifesting as clusters of co-morbidity. Patterns of multimorbidity affect individuals unevenly and have been associated with lifestyle, heritable traits and socioeconomic status^1–3^. Understanding each individual’s multi-morbidity risks is important to tailor healthcare decisions, motivate lifestyle changes or direct entrance into screening programs, as is the case for cancer^4,5^. Critically, health cannot only be understood by the presentation of individual diagnoses but rather in the context of an individual’s co-morbidities and their evolution over time. While a wide range of prediction algorithms exist for specific diseases from cardiovascular disease to cancer^6–8^, few algorithms are capable of predicting the full spectrum of human disease, which recognises more than 1,000 diagnoses at the top level of the ICD-10 (international classification of diseases, tenth revision) coding system.

Learning and predicting patterns of disease progression is also important in populations that are ageing and which exhibit shifts in their underlying demographic’s morbidities. For example, it has been predicted that globally, the number of cancer diagnoses will increase 77% by 2050^9^ or that in the UK, the number of working-age persons with major illnesses, including depression, asthma, diabetes, cardiovascular disease, cancer or dementia, will increase from 3 to 3.7 million by 2040^10^. Modelling the expected burden of disease is thus critical for healthcare and economic planning and, moreover, the continual tracking of disease occurrence along with its likely future prevalence within population groups promotes a more informed health system.

Recent developments in artificial intelligence (AI) may help overcome some methodological limitations of multi-morbidity modelling which have so far proved difficult to overcome^11^. Aside from the great number of diagnoses, these include challenges in modelling temporal dependencies among previous events, the integration of potentially diverse prognostically relevant data, and the statistical calibration of predictions. Large language models (LLMs)^12–15^, a subfield of AI that enables chatbots such as ChatGPT^16,17^, model language as a sequence of word fragments (so-called tokens). Generated token by token, the new text is based on all preceding text - and with enough training, the statistical dependencies among these tokens prove sufficient to produce context-aware and even conversational text, which is often indistinguishable from that of a human counterpart.

The analogy between large language models and disease progression modelling – which also entails recognising past events and exploiting their mutual dependencies to predict the future sequence of morbidity – has recently inspired a series of new AI models. For example, BERT-based models^18–21^ have been developed for specific prediction tasks. Transformer models trained on electronic health records have been used for predicting diseases such as pancreatic cancer^22^, self-harm^21^ and stroke^20^, as well as non-clinical parameters such as self-esteem^23^. However, despite promising proof of concepts^24^, the potential for comprehensive and generative multi-morbidity modelling has not yet been fully assessed.

Here, we demonstrate that attention-based transformer models, similar to LLMs, can be extended to learn lifetime health trajectories and accurately predict future disease rates for more than 1,000 diseases simultaneously based on prior health diagnosis, lifestyle factors and further informative data. Our extended model, termed Delphi-2M, is trained on data from 400,000 participants of the UK Biobank, a population-scale research cohort. The vocabulary of the model includes 1,256 ICD10 top-level codes, as well as sex, body mass, smoking, alcohol consumption and death. The analyses reveal that Delphi-2M predicts 97% of diagnoses with an average age and sex-stratified AUC (area under the receiver operating characteristic curve) of 0.76 (s.d. 0.08) for the next diagnosis and AUC of 0.73 (s.d. 0.08) for diagnoses 1 year into the future in internal validation data. Delphi-2M highlights how elements of past data influence the rates of subsequent diseases and enables sampling of future courses of health and disease at any point throughout an individual’s life course. Delphi performs similarly in longitudinal testing data from the UK (1-year gap average AUC 0.69, s.d. 0.09) and from Denmark without any additional finetuning (1-year gap AUC Denmark: 0.67 (s.d. 0.09). Delphi can provide competitive individual level prediction of multiple outcomes simultaneously and can model the overall expected evolution of a population from a specific health history starting point. In addition, the internal model of Delphi is partly interpretable and can provide a framework for future research into healthcare trajectories.

## Results

### A transformer model for health trajectories

A person’s health trajectory can be represented by a sequence of diagnoses using top-level ICD10 codes recorded at the age of first diagnosis as well as death. Further, “no event” padding tokens were randomly added at an average rate of 1/5 years to eliminate long intervals without other inputs, during which the baseline disease risk can significantly change (Extended Data Figure 1). Together, these data comprise 1,258 distinct states (or tokens in LLM terminology), each paired with the corresponding age, measured from birth, the sequence and timing of which are auto-regressively predicted. Additional health-related information includes sex, BMI, and smoking and alcohol consumption indicators, which are added at the age when the corresponding information becomes available but are not themselves part of predictions (**Figure 1a**).

**Figure 1.**
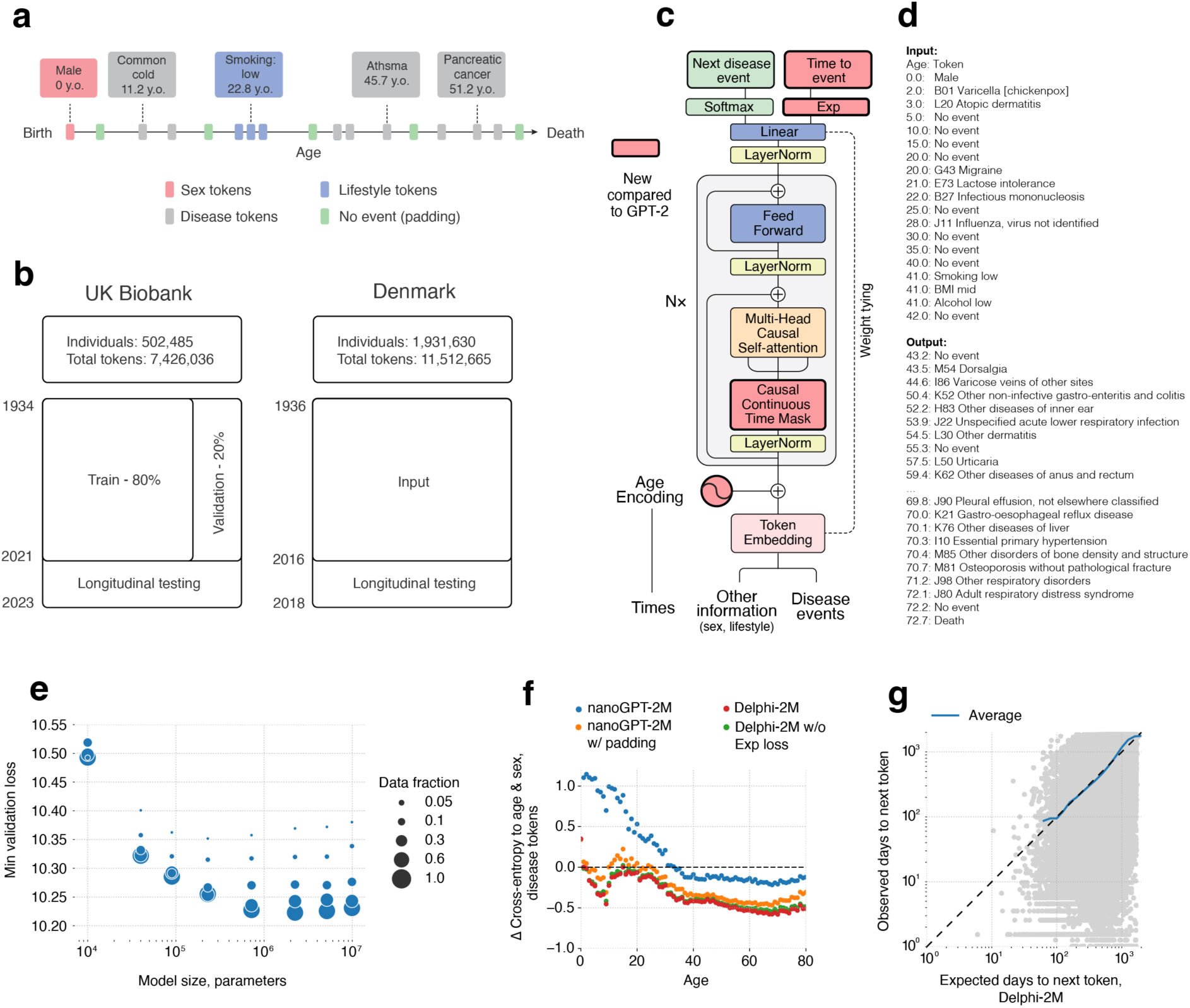
Delphi, a modified GPT architecture, models health trajectories. **a**, Schematic representation of health trajectories based on ICD-10 diagnoses, lifestyle and healthy padding tokens, each recorded at a distinct age. **b,** Training, validation, and testing data derived from the UK Biobank (left) and Danish disease registries (right). **c,** Delphi model architecture. Red elements indicate changes compared to the underlying GPT-2 model. **d,** Example model input (prompt) comprising (age: token) pairs and output. Input data is hypothetical and does not contain any real patient records; **e,** Scaling laws of Delphi showing the optimal validation loss as a function of model parameters for different training data sizes. **f,** Ablation results measured by the cross-entropy differences relative to an age-and-sex-based baseline (y-axis) for different ages (x-axes). **g,** Accuracy of predicted time-to-event. Shown are the observed (y-axis) and expected (x-axis) time to events for each next token prediction (grey dots). The blue line shows the average across consecutive bins of the x-axis.

Training data comprised 400,000 (80%) participants of the UK Biobank recorded before the 1st of July 2020. Data for the remaining 102,485 (20%) participants were used for validation and hyperparameter optimisation, while all records for 471,057 (94%) participants still alive on the 1st of July 2020 were used for longitudinal testing up until the 1st of July 2022 (**Figure 1b**). Additional external testing was carried out on the Danish disease registry data covering 1.93M Danish nationals aged 50-80 on 1. Jan 2016, with data available between 1978 and 2018.

To model disease history data, which, unlike text, occurs on a continuous time axis, we extended the GPT-2 architecture^25^ (**Figure 1c**). Transformer models map their inputs into a lower dimensional embedding space, in which information is successively aggregated to enable auto-regressive predictions. The first change thus replaces GPT’s positional encoding, a mapping that identifies each text token’s discrete position, with an encoding of continuous age using sine and cosine basis functions^12^. Standard GPT models only classify the next token using a multinomial probability model. Therefore, the second extension is the addition of another output head also to predict the time to the next token using an exponential waiting time model (**Methods**). As the rates of competing exponentially distributed waiting times define the logit probabilities of the next event, this parameterisation naturally extends GPT’s existing classification step (**Figure 1g**). Third, GPT’s so-called causal attention masks, which ensure that the model only accesses information from past events, are amended to resolve ties of tokens recorded at the same time (we note that such temporal ordering does not imply the model is capable of identifying causal mechanisms in the sense of modifiable features). Padding, lifestyle and sex tokens use a similar encoding but do not enter the likelihoods, as the model is deliberately not trained to predict them.

We term this model Delphi (**De**lphi **L**arge **P**redictive **H**ealth **I**nference). This architecture allows one to provide the model with a partial health trajectory (prompt in LLM terminology) in order to calculate the subsequent rate (per day) for each of the 1,256 disease tokens plus death. Furthermore, the next token and the time to this event can be sampled based on these rates. Iteratively, this procedure samples entire health trajectories (**Figure 1d**).

A systematic screen of architecture hyperparameters (embedding dimensionality, number of layers, heads) confirms the reported empirical scaling laws^26^, which state that model performance increases with the number of data points and, up to a limit defined by the available data, as the number of parameters increase (**Figure 1e; Extended Data** Figure 2). The screen indicates that for the UK Biobank dataset, optimal Delphi models have around 2M parameters. One of the models within the optimal range, contains 120 internal embedding dimensions, 12 layers and 12 heads, amounting to a total of 2.2M parameters. Results based on this model parameterisation are discussed throughout the rest of the paper. We note that qualitatively similar results are obtained from other parameter choices (Extended Data Figure 3).

An ablation analysis shows how Delphi-2M’s architectural modifications contribute to a better age-and-sex-stratified cross-entropy compared to a standard GPT model (**Figure 1f**; **Table 1; Extended Data** Figure 4). A good, albeit slightly inferior, classification performance at different ages may already be achieved by adding regular “no event” padding tokens to the input data with GPT models alone. However, a key distinguishing feature of Delphi compared to basic GPT models is its ability to calculate the absolute rates of tokens, which provide consistent estimates of inter-event times (**Figure 1g**). This property also implies that the rates may be interpreted as the incidences of tokens, as recorded in the training data.

**Table 1.** Ablation results for Delphi-2M. Top. Change in cross-entropy of disease token predictions relative to a baseline derived from age (in years) and sex incidence. **Bottom.** Change in the exponential loss for the waiting time until the next token.

| Loss | Age and Sex | GPT | + healthy tokens | + age enc. & masks | + time loss |
| --- | --- | --- | --- | --- | --- |
| Next token class (cross-entropy) | 0 (baseline) | −0.206 | −0.478 | −0.567 | **−0.574** |
| Time to next token (exponential) | 0 (baseline) | N/A | N/A | N/A | **−0.19** |

### Delphi-2M accurately models reported multi-disease incidences

Delphi-2M’s accuracy in predicting diverse disease outcomes in the validation cohort is compared to the sex and age-stratified incidence as an epidemiological baseline. As can be seen in the ten examples shown in Figure 2a, the incidence curves are very varied, with some diseases, such as chickenpox, peaking in infancy, whereas others, such as asthma or depression, are relatively flat and with most rising exponentially in old age. Moreover, there are noticeable differences between the sexes, which are obvious for breast cancer but also noticeable for diabetes, depression, acute myocardial infarction and death. Delphi-2M’s predictions are updated for each individual when new inputs are recorded. The predictions largely follow the sex- and age-stratified incidence curves but also indicate events or periods when the individual risk remains below or rises above the population average. For some diseases, such as asthma or arthrosis, the spread is narrow, indicating a limited ability to predict beyond the sex- and age-incidence trend. Yet for other diseases, including septicaemia, and also death, the spread is wide, indicating measurable inter-individual differences in disease rates.

**Figure 2.**
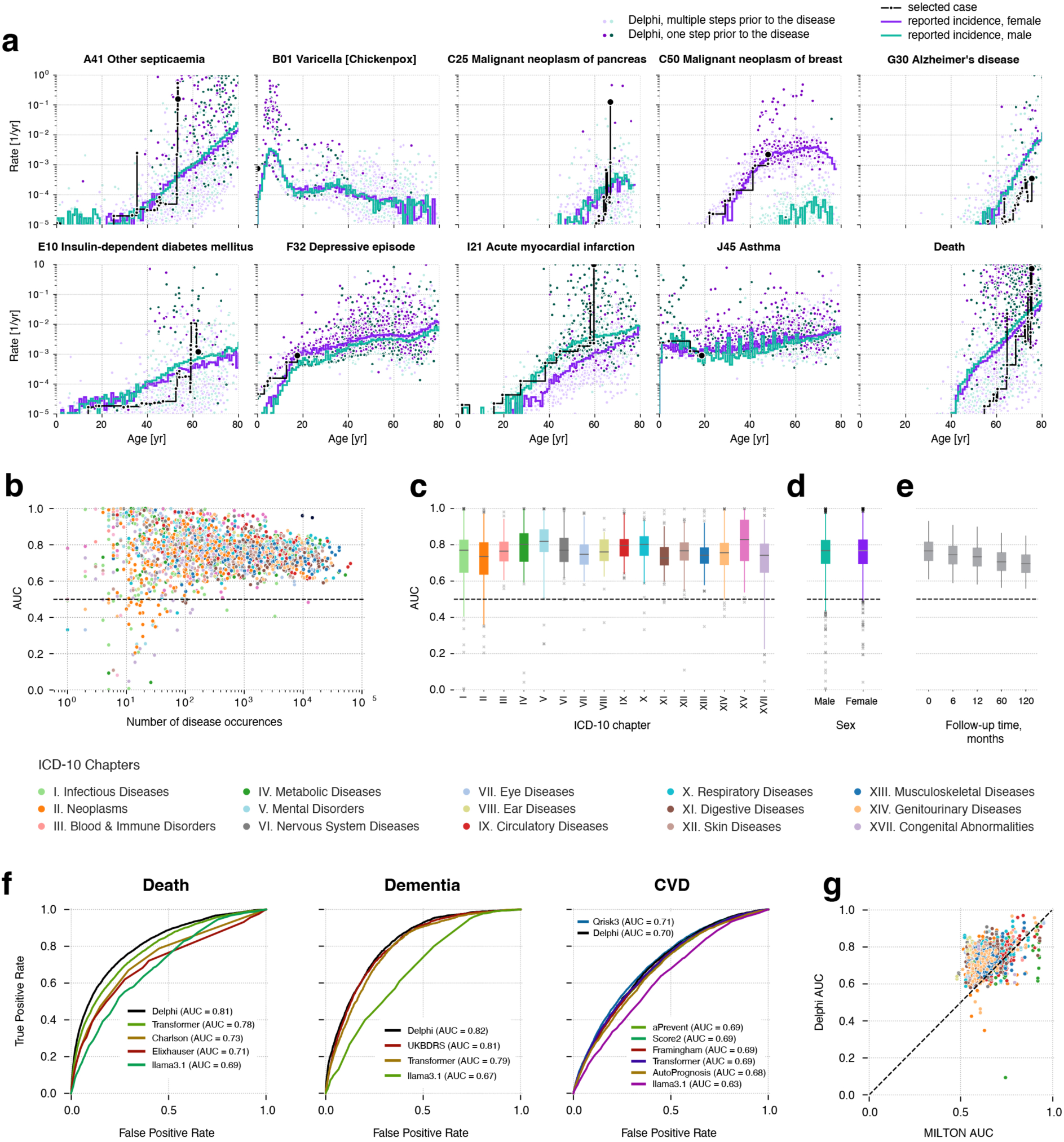
Delphi-2M accurately models the rates of a wide range of diseases. **a**, Predicted rates for 9 exemplary diagnoses and death (y-axis) shown as a function of age (x-axis). Points show predictions at each recorded input token. Colours separate biological sex; the darker colours indicate predictions immediately prior to the diagnosis in question. The purple and turquoise lines are disease rates observed for each yearly age bin in the training data. The solid black line connects consecutive predictions for one randomly selected case throughout age. **b**, Average age-sex-stratified AUCs (y-axis) as a function of training occurrences (x-axis). Shown are data for n=906 diagnoses for males and n=957 diagnoses for females for which sufficiently many events were recorded in the validation data to evaluate AUCs. **c**, Same as **b**, aggregated by the ICD-10 chapter. The boxplots feature the median as the center line, the box from the first to the third quartile and the whiskers for 0.025 and 0.975 quantiles. **d**, Same as **b**, aggregated by sex. The boxplots feature the median as the center line, the box from the first to the third quartile and the whiskers for 0.025 and 0.975 quantiles. **e**, AUCs of all diagnoses in **b** for different time gaps between prediction and diagnoses (x-axis). **f,** ROC curves for Delphi and other clinical or machine learning methods for three selected endpoints evaluated on the internal longitudinal testing set. **g,** AUCs of MILTON^27^, a biomarker-based machine learning model (x-axis) in prognostic mode compared to Delphi-2M AUC values from the UK Biobank validation set (y-axis) for n=410 diagnoses.

Delphi’s ability to predict the next diagnosis token across the spectrum of human disease is confirmed by the average age-stratified AUCs (area under the receiver operating characteristic), which averages at values of approximately 0.76 in the internal validation data (Figure 2b**; Extended Data Table 1**). For 97% of diagnoses, the AUC was greater than 0.5, indicating that the vast majority followed patterns with at least partial predictability. These patterns were found to be true across the different chapters of the ICD-10 spectrum, which define broad groups of disease for both sexes (Figure 2c**,d**). Among the most confidently predicted next events is death, with an age-stratified AUC of 0.97 in both sexes. Importantly, calibration analyses in 5-year age brackets show that the predicted rates closely match the observed number of cases, showing that the models’ rates of the next tokens are consistently estimated (Extended Data Figure 5).

Next event predictions are often the consequence of acute illness or diagnostic refinements that accrue over the course of a few weeks or months, which may be undesirable for prognostication. Delphi-2M’s average AUCs decrease from an average of 0.76 to 0.70 after 10 years, indicating that its predictions are also relevant for prognostication (Figure 2e**, Extended Data** Figure 6). Similar results were observed in longitudinal test data, which also shows that there was no substantial shift in diagnostic patterns throughout Biobank’s follow-up (Extended Data Figure 7).

Delphi-2M performs equally, or better than existing risk scores, machine learning models and LLMs for cardiovascular disease, dementia and death (Figure 2f**, Extended Data** Figure 7c**, Extended Data Table 2**). This was the case for next event predictions, as well as prediction horizons up to 24 months. Delphi-2M’s AUCs were also generally higher than those of a recent machine learning algorithm which calculates the risks of a similarly broad spectrum of ICD-10 diagnoses using 67 different biomarkers available through the UK Biobank^27^, even though for many diagnoses, such as diabetes, biomarkers remain indispensable (Figure 2e**; Extended Data** Figure 8). This shows that Delphi-2M’s multi-disease predictions match or exceed current risk models for individual disease outcomes and offer the great advantage of enabling the simultaneous assessment of more than 1,000 diseases at any given time while also surpassing multi-disease models in quality.

### Generative modelling of future disease trajectories

One of the most promising features of generative models is the ability to sample disease trajectories, conditional on data recorded up to a certain point. This is a property which few conventional epidemiological models possess.

In order to systematically assess the influence of medical histories on future health, we sampled health trajectories for each participant from the UK Biobank validation cohort based on data available until the age of 60 (Figure 3a). This provides the opportunity to compare 63,662 sampled and observed trajectories. When evaluated at the population level, the disease incidences at ages 70-75 are well recapitulated, showing that the overall distributions are well preserved by iterative sampling (Figure 3b). This is further confirmed by the cross-entropy loss of sampled trajectories, which is, on average, indistinguishable from the observed data but drops when the preceding disease histories are shuffled between participants (Extended Data Figure 9a,b).

**Figure 3.**
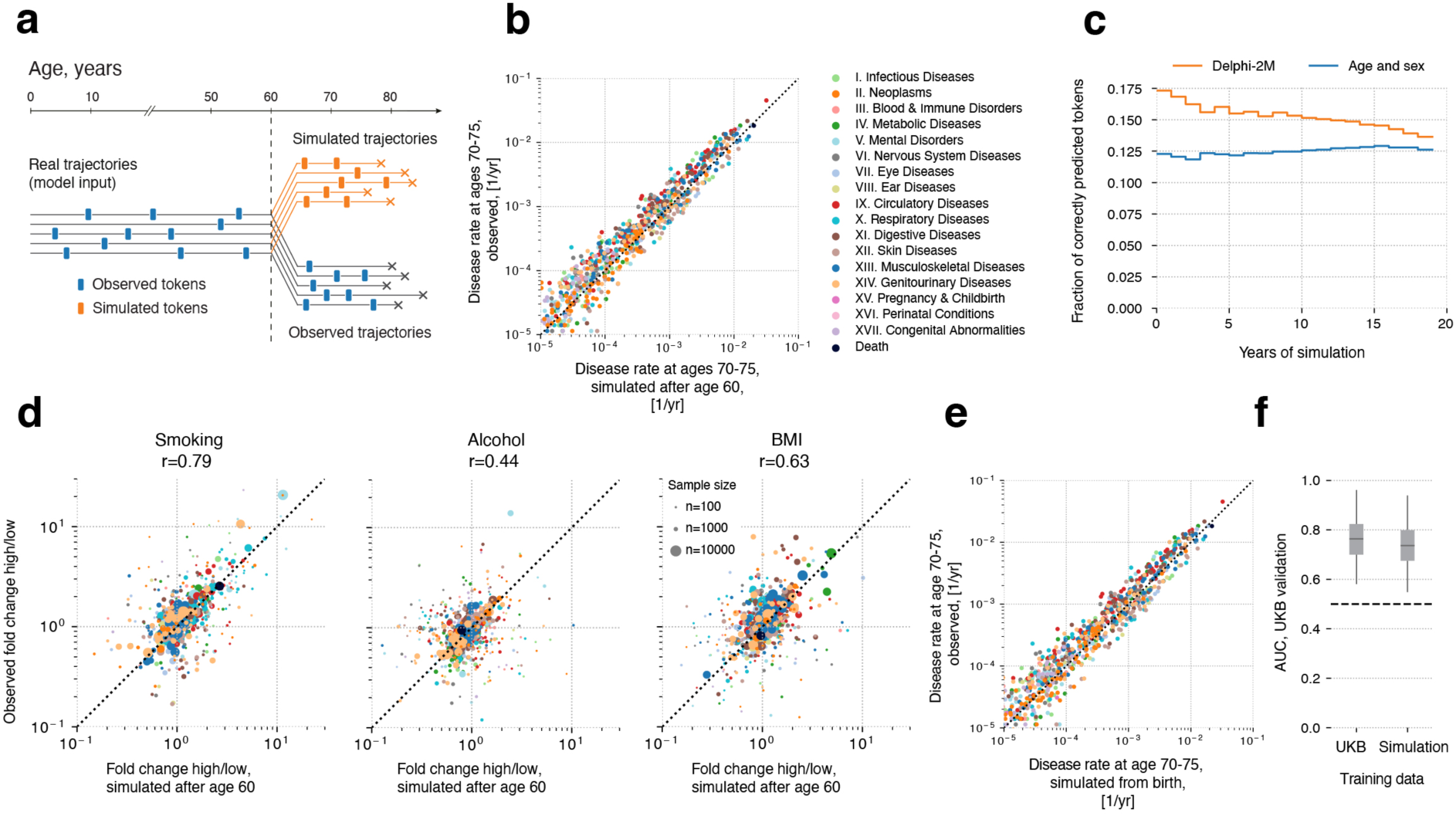
Generative modelling with Delphi-2M informs future outcomes. **a**, Schematic representation of the experiment design. Delphi-2M is used to simulate health trajectories using validation data (n=100,000 individuals) observed until the age of 60. A single trajectory is simulated per individual. Subsequently, simulated trajectories are compared to the observed outcomes for the same person. **b,** Delphi-2M-modelled disease rates at ages 70-75 (x-axis) compared to observed rates at the same ages (y-axis). **c,** Fraction of correctly predicted diagnoses (y-axis) per 1-year age bin as a function of the years after simulation started at age 60 (x-axis). Delphi-2M, orange. The blue curve uses age & sex as a prediction baseline. **d,** Simulated (x-axis) and observed (y-axis) fold changes of disease rates for high versus low smoking, alcohol consumption and BMI groups. The evaluation period included ages 70-75 and used simulations from the age of 60. **e,** Same as **b,** shown evaluated for simulations from birth. **f.** Boxplots for AUCs of disease risk prediction for Delphi when trained on UKB and Delphi-2M-sampled synthetic data (**Methods**).

In the first year of sampling, there are, on average 17% disease tokens that are correctly predicted, which drops to less than 14% 20 years later. These figures compare to values of 12-13% of correctly predicted disease tokens using sex and age alone, confirming that the conditional generation helps make more accurate predictions of future events (Figure 3c**, Extended Data** Figure 9c,d).

Delphi-2M’s ability to simulate differential health outcomes over a decade or more, based on each individual’s health history, manifests in a multitude of ways. For example, the changes in disease burden in different population subsets defined by smoking, alcohol consumption or BMI are well predicted (Figure 3d**, Extended Data** Figure 10a). Similar findings are observed when the population is stratified by the presence of prior diseases or by estimated disease risks (Extended Data Figure 10b,c). Together, these analyses show that Delphi-2M’s conditional samples provide meaningful extrapolations for future health courses, which reflect the influence of past health events.

The use of synthetic data has been proposed to help overcome issues with privacy in biomedical modelling if such data sets do not reveal characteristics specific to any one person. Fully synthetic data, which is sampled from birth with randomly assigned sex, reproduce the observed age and sex-specific incidence patterns throughout life (Figure 3e**; Extended Data** Figure 8). Further assessment shows that the generated trajectories do not exhibit any greater similarity to the training data than those from the validation cohort (Extended Data Figure 11). While partially overlapping disease trajectories may be found in terms of absolute disease tokens, the extent of overlap appears as expected based on the observed incidences and co-morbidity patterns.

To illustrate the utility of synthetic data, we trained a version of Delphi-2M exclusively on synthetic data. Remarkably, when evaluated on the observed validation data, the fully synthetically trained model achieves an age-sex stratified average AUC of 0.74, which is only three percentage points lower than that of the original Delphi-2M model (Figure 3e; Extended Data Figure 9g). This confirms that synthetic data preserve much of the information relevant to training Delphi models and may serve as a less privacy-sensitive alternative to personal data.

### Explainable AI provides insights into disease progression

Insights into how Delphi-2M utilises past information to predict future disease rates can be obtained by assessing the structure of the disease embeddings. GPT models linearly map inputs into a lower dimensional embedding space, in which the temporal sequence of events is iteratively aggregated to produce a state from which predictions of each next token are derived (Figure 1c). Delphi-2M’s specific implementation uses the same mapping from this final embedding state to the token risk, which guides the interpretation as the embedding matrix reflects the observed structure of co-morbidity risks.

As shown by the UMAP representation of Delphi-2M’s embedding matrix in Figure 4a, disease codes cluster closely by the underlying chapter, a property the model has no direct knowledge of, and which purely reflects co-occurrence patterns in the data. Yet there are also noticeable exceptions, for example, cancers and precancers of the female reproductive tract. Another noteworthy cluster involves the two types of diabetes, retinal disorders and the neuropathies caused by them. Diseases with high acute mortality, such as myocardial infarction or septicaemia, are clustering with death.

**Figure 4.**
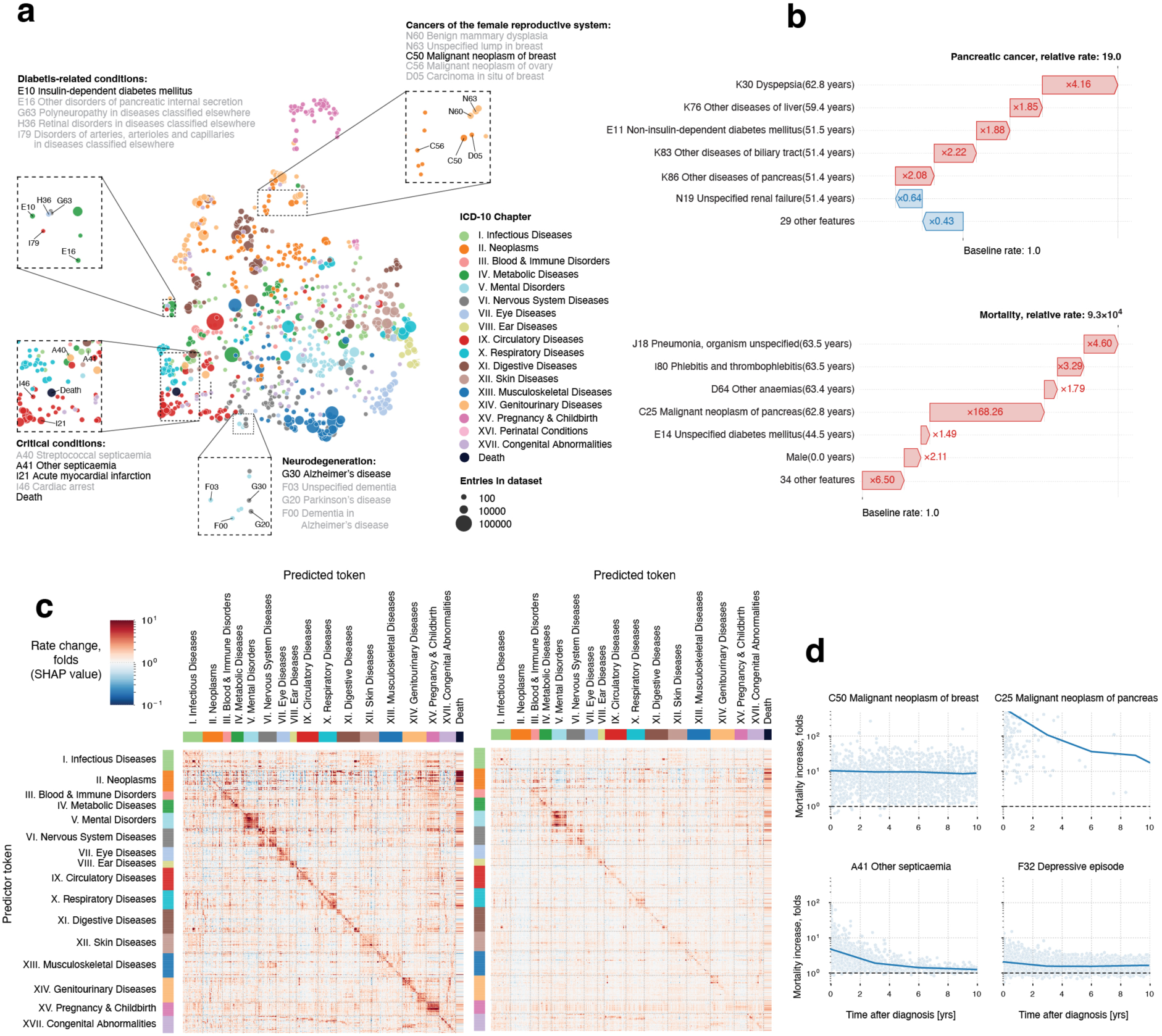
Explainable AI offers insights into disease progression. **a**, UMAP projection of token embeddings. Selected diseases are shown in the zoomed-in areas. Colours define disease chapters. **b,** SHAP-explained token risk contributions for individual trajectories. The top shows the risk of pancreatic cancer immediately prior to diagnosis at age 68.2, which was found to be 19x increased. The bottom panel shows the SHAP estimates of contributions to estimated mortality at age 63.5, which was greatly increased, in large part due to the preceding diagnosis of pancreatic cancer. **c,** Heatmap of average SHAP effect of each of n=778 disease tokens with more than 5 occurrences and grouped by chapter (y-axis) on the same set of tokens plus death (x-axis). Red colours indicate a risk increase, whereas blue is a decrease. **d,** Rate change (SHAP value) of mortality (y-axis) as a function of time after diagnosis (x-axis) for selected diseases.

To gain further insights into how individual tokens influence future risks SHAP (SHapley Additive exPlanations)^28^ values measure the influence of each token from the input trajectory on model predictions by a systematic assessment of subsampled data for individual prediction. As illustrated with the example of the proband’s partial trajectory shown in Figure 4b, this analysis reveals that a series of disease diagnoses of the digestive tract (Chapter XI) elevated their pancreatic cancer risk 19-fold. The subsequent pancreatic cancer diagnosis, in turn, increased the rate of mortality almost ten thousandfold.

SHAP analysis of data from 100,000 individuals of the validation cohort reveals the mutual dependencies by which each disease, sex and lifestyle token influences the rate of subsequent disease tokens, similar to hazard ratios in conventional statistical models (Figure 4c**, left; Extended Data** Figure 12). Effects mostly increase the rates of other diseases and are usually found among diseases of the same chapters, underscoring that the recorded patterns of co-morbidities often cluster within specific ICD-10 disease chapters. Particular clusters spanning entire disease chapters are visible for chapters V. Mental Disorders and XV. Pregnancy & Childbirth. Notably, such patterns often appeared symmetrical, indicating similar predicted effect sizes of one disease token influencing another and vice versa (Extended Data Figure 13). This behaviour can be attributed to the structure of the embedding space, which places temporarily co-occurring diagnoses in local proximity (Extended Data Figure 14). Due to the linear relation between embedding and logarithmic disease rates local proximity in the embedding space also leads to a similarity of modelled disease rates.

The patterns of modelled influences after 10 years are similar to short-term effects, even though the strength of associations is greatly attenuated (Figure 4c**, right)**. The cluster relating diseases of Chapter XV. Pregnancy & Childbirth within 5 years is entirely absent after 10 years, which is expected given that pregnancy-associated diseases occur within a finite period. Dependencies among mental disorders, however, remain apparent, similar to the effects of neoplasms on mortality. These observations are noteworthy as the quantification of temporal dependencies on past events poses a particular challenge for conventional epidemiological models, whereas Delphi-2M’s GPT model uses attention-based weights, which are updated with every new input, including the ‘no event’ paddings.

To further illustrate Delphi-2M’s capabilities of modelling temporal dependencies, we note that for some diseases, such as cancers, the influence on mortality decays with a half-life of several years, reflecting the sustained risks of recurrence or impacts of treatment (Figure 4d). For septicaemia, however, the influence on mortality is much more short-lived and drops sharply, effectively recovering to values close to the population average. This inference agrees with traditional Nelson-Aalen analyses of the hazard rates (Extended Data Figure 13f). This behaviour is also reflected by Delphi-2M’s attention maps, which show that cancer tokens are attended to for long periods, while those of septicaemia, myocardial infarction and many other diseases tend to be short-lived (Extended Data Figure 15).

### External testing using Danish disease registries

The previous sections showed that Delphi-2M accurately reproduces health trajectories and the underlying complex dependencies of health and disease tokens as recorded in the UK Biobank. However, there is a risk that such patterns are specific to the particular training data and do not generalise to data sets from other populations. While the UK Biobank cohort has been recruited according to epidemiological best practices, due to the age of recruitment and the volunteer nature, the demographics are not entirely representative of the general population of the UK^29^. Furthermore, there may be differences in the availability of health-related data and how it is recorded between different healthcare systems.

To assess whether Delphi-2M’s inference generalises to data from another country, we performed external testing using Danish registry data. For this purpose, we transferred Delphi-2M with the weights learned from the UK Biobank training and evaluated predictions on the Danish data; hence, no retraining or adjustments have been made. We use a cross-section of the entire Danish population aged 50-80 years on 1. Jan 2016 and evaluate Delphi-2M’s predictions on the incidence of the corresponding population between 1. Jan 2017 and 1. Jan 2018, therefore, enforcing a 1-year data gap to also assess prognostication. Data are collected for 1.93M individuals (51% females and 49% males), with 11.51M disease tokens recorded between 1978 and 2016. Predictions are evaluated on 0.96M disease tokens across 796 ICD10 codes (each with at least 25 cases).

The average AUC when Delphi-2M is applied to Danish data was 0.67 (s.d. 0.09) with similar performance across most chapters. The AUC difference to age-sex baselines was on average 0.07 (s.d. 0.08) points higher for Delphi-2M, with 83% of all diseases showing a signal above the baseline (Figure 5a). When compared to the longitudinal testing in the UK Biobank, AUC values are similar, with a correlation of 0.76 (95% CI: 0.72–0.80) (Figure 5b). Further, the AUC differences over an age-sex baseline remain comparable across the two cohorts with a correlation of 0.54 (95% CI: 0.47–0.60) (Figure 5c).

**Figure 5.**
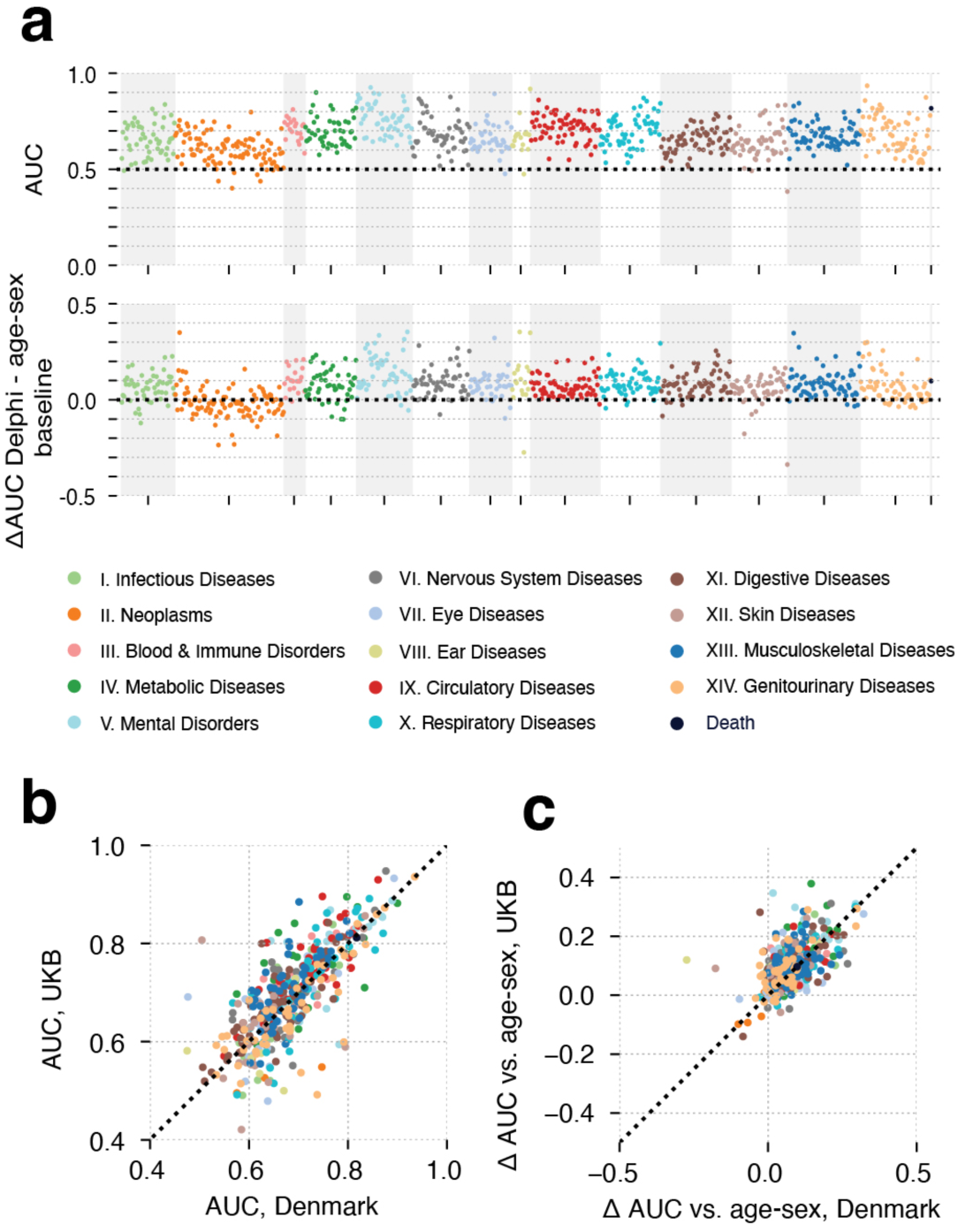
External validation using Danish registries. **a**, AUC results of Delphi for each token with at least 25 occurrences coloured by the respective ICD-10 chapter. Predictions are based on Danish adults (50-80 years of age) with data up until 1. Jan. 2016 and evaluated on incidence between 1. Jan. 2017 - 1. Jan. 2018. Below are the respective differences in AUC values between Delphi and an age-sex baseline based on Danish data from 2010-2016. b, Comparison between delta AUC values in the UKB longitudinal testing and the Danish testing. c, Comparison between AUC values in the UKB longitudinal testing and the Danish testing with their respective age-sex baselines.

Individual predictions of specific disease risks recapitulate changes of up to three orders of magnitude across the population and are slightly less calibrated than predictions of UK Biobank participants (Extended Data Figure 5b). Especially for mortality a systematic underestimation is observed, which we will discuss in the next section. Overall, Delphi-2M’s predictions transfer well across two healthcare systems, without any reconfiguration, and show considerable prognostic performance across most diseases.

### Limitations, biases and fairness

While AI models can learn diverse and complex patterns, they may also replicate biases that exist in the underlying training data. Delphi-2M is no exception to this and its reduced accuracy on Danish registries suggests a range of biases. In particular, we identify and discuss the consequences of the following biases underlying UK Biobank training data: Recruitment bias, sampling/immortality bias, ancestry background, levels of deprivation and health data availability bias. Similar limitations will exist to some degree in most other data sources and are important considerations when training AI models on healthcare data.

Epidemiological cohorts often only approximately reflect the general population due to differential response rates in different subpopulations. The UK Biobank comprises more white British citizens than other ethnicities, and participants tend to be, on average, more affluent and educated than the general population^29^. Other biases arise from the use of retrospective data in a cohort. In the context of the UK Biobank, most individuals have been recruited between 40-70 years of age. This creates a type of selection bias as no deaths are recorded prior to recruitment (e.g. conditional on having survived until recruitment). This has direct implications on estimated mortality (Figure 6a) and indirectly also on the incidences of diseases associated with high mortality, such as cancers, as only survivors are included in the UK Biobank. For time-dependent analyses, the jump of mortality to non-zero values at recruitment can also lead to false attribution of the apparent increase to unrelated variables recorded at the time of recruitment. Furthermore, limited follow-up data are currently available for ages greater than 80 years. This period is, therefore, not reliably modelled by Delphi-2M.

**Figure 6.**
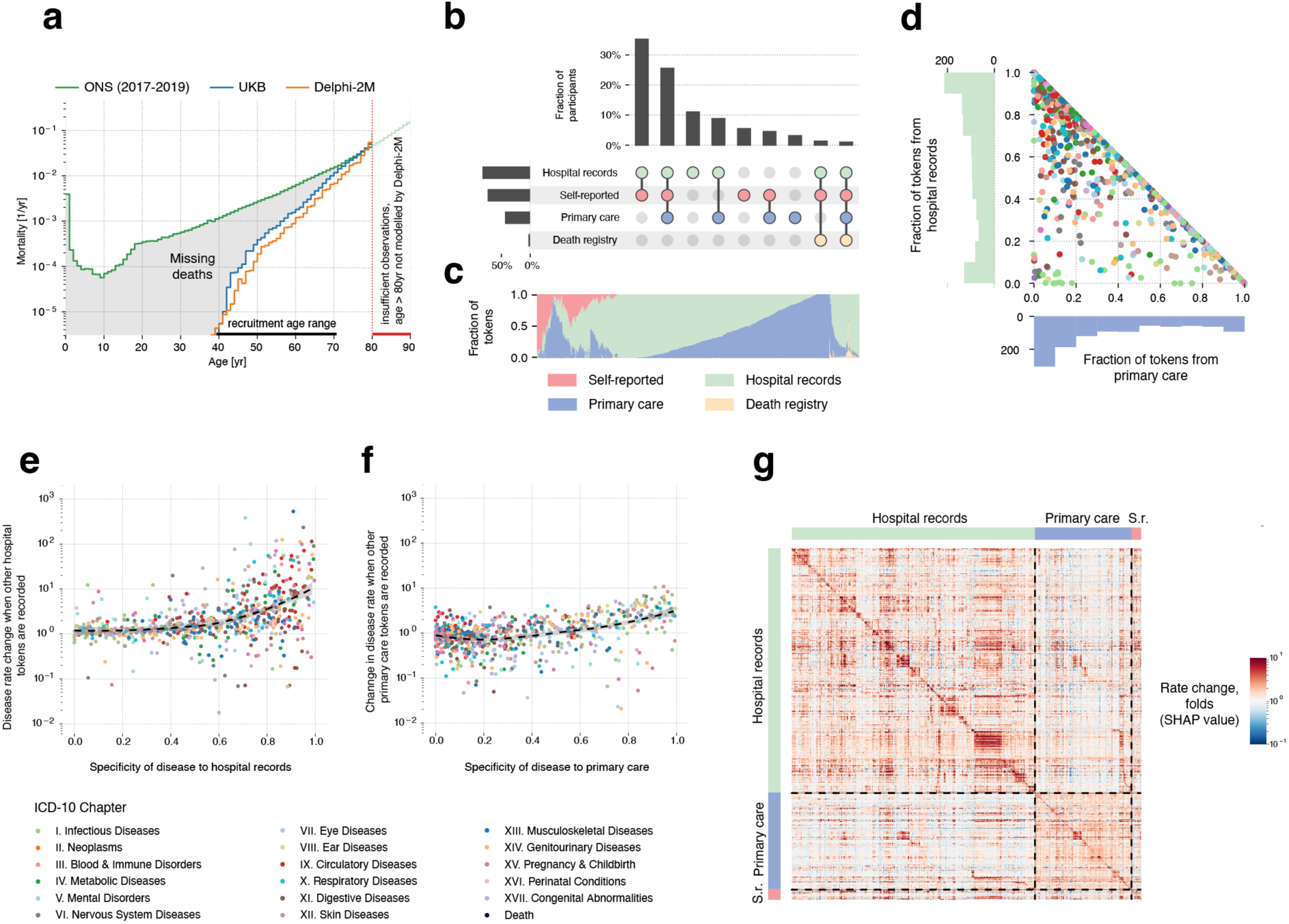
Epidemiological biases in UK Biobank data reflected by Delphi-2M. **a,** Yearly mortality estimates by Delphi-2M (UK validation cohort), observed rates in the UK Biobank and ONS national estimates across the entire British population. As only living individuals between 40-70 years of age (black line) were recruited to the UK Biobank, many deaths are missing compared to the ONS population estimate (grey shaded area). **b,** UpSet plot of disease data availability in UK Biobank validation cohort (n=100,000). **c,** Data source distribution for records per disease token (token position sorted). **d,** Records source fractions for two main sources, for all diseases, coloured by the ICD-10 chapter. **e,** Hospital-records missingness bias (relative rate after first hospital token; y-axis) as a function of token exclusivity to hospital records (y-axis) for each relevant diagnosis (points). Points are coloured by the ICD-10 chapter, the overall trend is shown in black using a non-parametric (loess) curve with 95% confidence intervals shown in grey (UK validation cohort). **f,** Primary care missingness bias (y-axis) as a function of primary care token exclusivity, coloured by the ICD-10 chapter. The trend is shown in black using a non-parametric (loess) curve with 95% confidence intervals shown in grey (UK validation cohort). **g**, SHAP value matrix, similar to Figure 4c. Columns and rows correspond to different diseases and are sorted by the dominating source, then ICD-10 chapter. A dominating source is defined as the origin of more than 65% of records for a given disease; diseases without a dominating source are not shown. SHAP values indicate the greater influence of diseases on other diseases from the same group.

Furthermore, the heterogeneous sources of UK Biobank’s first occurrences data and their missingness patterns introduce artificial correlations. The first occurrence data has been collated from self-reports, primary care, hospital admissions, cancer and death registries, which were only available for different overlapping subsets of participants and time periods (Figure 6b**,c****; Extended Data** Figure 16). Moreover, each source contributes characteristic disease tokens. Self-reporting and GP records contain mostly common diseases, with self-reporting covering 78% of participants, while GP records were only available for just 45%. Data from hospital records covered 86% of participants and contributed more aggressive disease tokens, such as myocardial infarction or septicaemia, similar to the death registry. Lastly, cancers were provided by the national cancer registry. Many diagnoses exhibit a distinct source specificity (Figure 6c**,d**).

Delphi-2M can learn patterns of missingness and use them to predict future rates accordingly – even though this concerns, in many cases, data availability rather than true incidence. The predicted rates of diseases exclusive to hospital records are, on average, 10x higher in individuals with a history including other hospital records (Figure 6e). Septicaemia, for example, is diagnosed in 93% of cases in a hospital setting and is predicted to occur at 8x greater rates in individuals with any other hospital data. Similarly, the predicted rates of diseases exclusive to primary care, such as acute sinusitis, are, on average, 3x higher in participants with other primary care data (Figure 6f). While some of these associations may reflect true diagnostic pathways or disease clusters diagnosed in a distinct care setting, it nevertheless appears that many of these associations are artefacts stemming from the incomplete aggregated nature of UK Biobank’s data. These source effects also explain some of the substructures visible in the UMAP representation of disease embeddings (Figure 4a**, Extended Data** Figure 17a-d) and also in the matrix of SHAP effects (Figure 4c**, 6g; Extended Data** Figure 17).

An important consideration of healthcare models is whether predictions are made with similar accuracy in different population groups. The predicted rates of observed diseases are generally comparable between the sexes (Extended Data Figure 18). Across self-reported ethnic backgrounds, however, lower rates for individuals self-reporting “white” are predicted, reflecting the observed disease rates (Extended Data Figure 18a,c). Within each ethnicity, Delphi-2M exhibits similar AUCs, with a trend towards lower values in the white ethnicity group (Extended Data Figure 18g). Similarly, Delphi-2M’s predicted rate of disease is elevated for groups with a higher Townsend deprivation index, which mirrors the average number of tokens per individual in this group and coincides with a higher AUC (Extended Data Figure 18b,d,f). Simulated trajectories are also found to reproduce the observed disease burden in these groups (Extended Data Figure 19). Together these observations indicate that Delphi-2M’s predictions reflect the differential burden of disease seen across different population subgroups but exhibit mostly comparable discriminatory performance within each stratum.

## Discussion

Here, we presented Delphi-2M, a GPT-based prototype model of multi-disease progression. Delphi-2M extends the GPT large language model to account for the temporal nature of health trajectories. Analogous to LLMs, which learn the grammar and contextual logic of language from large bodies of text, Delphi-2M inferred the patterns of multi-disease progression when trained on data for more than 1,000 diseases and baseline health information recorded in 400k UK Biobank participants.

A detailed assessment of Delphi-2M’s predictions showed that they consistently recapitulate the patterns of disease occurrence at the population scale as recorded in the UK Biobank. For the majority of diseases, Delphi-2M’s multi-disease, continuous-time model predicted future rates at comparable or better accuracy as established single-disease risk models, alternative machine learning frameworks and also blood biomarker based models. Only a small performance drop was observed when applied to data from Danish disease registries, demonstrating that models are — even without additional finetuning — largely applicable across national healthcare systems.

Delphi-2M is uniquely capable of sampling future disease trajectories, which enables estimating cumulative disease burdens over periods of up to 20 years, conditional on prior health information. It is worth noting that Delphi’s predictions are generally strongly influenced by statistical chance and compatible with a range of outcomes for a given individual. The ability to generate synthetic data may also help create data sets that preserve the statistical co-occurrence patterns without revealing any specific data, which could facilitate the development of further AI models with a decreased risk of revealing personal information.

Delphi-2M’s underlying GPT-2 model offers insights into the modes of disease progression. The ability to cluster disease risks may be useful for genomic association studies that focus on comorbidities or are stratified by the risks derived from health trajectories. Delphi-2M’s capability to quantify the temporal influence of previous health data revealed that cancers increase mortality in a sustained fashion, while the effects of myocardial infarction or septicaemia regressed within 5 years. Similar analyses also revealed clusters of persisting comorbidities, such as mental health conditions, which informs healthcare planning. While Delphi-2M appears capable of modelling temporally directed dependencies, we caution, however, against interpreting these as causal relationships that could be exploited to modify future health courses.

There are also a range of limitations that need to be considered. A detailed analysis of UK Biobank’s first occurrence data revealed several biases, reflected by Delphi-2M. In addition to a healthy volunteer bias and selection bias of participants before recruitment, the diverse nature of health data sources impacted Delphi’s prediction as UK-Biobank-specific patterns of missingness are exploited to infer disease rates. Furthermore, Delphi-2M predicts different disease rates in subgroups based on ancestry background and deprivation indices but no observable trend between lifestyle measures and birth year. These findings underscore that care must be taken when AI models are used for inference and applied for prediction in heterogeneous healthcare data sets.

A promising feature of Delphi-2M’s implementation is the relative simplicity with which additional data layers may be incorporated in future extensions. Examples of this are the simple additive nature of age encoding or how lifestyle and sex tokens are superimposed on the embeddings of disease history information. Immediate refinements of Delphi-2M may incorporate additional lifestyle data, prescription records and blood tests, both usually available in a general healthcare setting. Further multi-modal extensions could include genomic data, richer metabolomic information, or data from wearables that can be added to Delphi-2M’s embedding layer, similar to how lifestyle tokens are currently incorporated. Such multi-modal extensions will shed light on how different layers of biology interface in shaping health outcomes and thus help address long-standing questions in biomedicine. Furthermore, while ICD-10 provides a predefined tokenisation of diseases, LLMs have been shown to also conceptualise natural language, making it plausible to expect that future models may derive similar meaning directly from free text records, enabling applying Delphi-like models to unstructured data. Lastly, Delphi-2M itself could serve as an extension to LLMs. Similarly to systems that provide LLMs with query-relevant web search results to reduce hallucinations^30^, a future healthcare-oriented LLM could invoke a Delphi-based model to improve the numerical accuracy of the generated replies^31^.

Currently, Delphi-2M constitutes a proof of concept that AI architectures originally developed for natural language processing can be extended to model the temporal sequences of complex healthcare events. Notwithstanding Delphi-2M’s current limitations, its encouraging performance relative to other methods and its transferability between UK and Danish healthcare systems indicated there will be clinical utility of Delphi or similar tools in the future.

An evident application of Delphi-type models would be to support medical decision-making by the probability distribution of future trajectories for a patient. A direct use could be to identify individuals who meet established risk thresholds, e.g. by age or biomarkers, that warrant inclusion in screening programs or further prevention measures. As discussed above, it would be critical to assess and mitigate biases present in the training set before any such system is tested in a clinical setting. Even with a relatively unbiased model, the benefits of such risk stratification must first be assessed through randomised controlled trials before broader implementation. Additionally, deploying clinical decision support systems requires a regulatory framework, which is still in its infancy for AI in healthcare. A long-term objective for AI-based clinical decision support systems is to provide direct treatment recommendations. Currently, Delphi does not explicitly model the effect of treatments, and usually, it is only possible to estimate these effects reliably using data from randomised clinical trials. Therefore, it is essential to establish frameworks within which clinical trial data can inform AI models, which ar,e in turn, to be evaluated in subsequent randomised studies.

An alternative utility of Delphi models would be to help allocate healthcare resources by identifying the needs for future interventions. At a local level, for example, this could inform care trajectories on admission to a hospital. Such applications, informing the providers rather than treatment decisions, require less regulation. Similarly, there may be substantial system-wide modelling benefits for Delphi type models, in particular in the tailored projection of a cohort using their current healthcare until a point in time. This has the potential to produce better estimates of the future for aggregate healthcare demand, which is useful at regional and national health system levels, as well as for insurers. This will help allocate resources that can be scarce in ageing populations.

These considerations illustrate the wide range of applications of generative models for biomedical research and, ultimately, also for healthcare. With appropriate training and evaluation, future multi-modal model extensions may be used for preventative medicine, clinical decision support and healthcare planning. Our model and analysis presented here present a further step to unlock the considerable healthcare benefits from the era of AI.

## Methods

### Data

#### UK Biobank

##### Cohort

The UK Biobank is a cohort-based prospective study comprising approximately 500,000 individuals from various demographic backgrounds recruited across the UK between 2006 and 2010. At the time of recruitment, individuals were between the ages of 37 and 73^32^.

##### Disease first occurrence data

The main data source for health-related outcomes is built on the first occurrence data assembled in category 1712 from the UK Biobank. These data include ICD-10 level 3 codes (e.g. E11: Type 2 diabetes mellitus) for diseases in chapters I-XVII excluding chapter II - neoplasms (1256 in total) plus death. The data is pre-assembled by UK Biobank and includes the first reported occurrence of a disease in either the linked primary care data (cat. 3000), inpatient hospital admissions (cat. 2000), death registry (fields: 40001, 40002) or self-reported data through questionnaires (field: 20002).

Information on neoplasms was not included in category 1712 by the UK Biobank, hence we included the data ourselves through the addition of the linked cancer registry data in fields 40005 and 40006 (subset to the first occurrence and mapped to ICD10 level 3 codes).

##### Lifestyle and demographics

We extract information on the self-reported sex of participants as recorded in field 31 (indicators for female and male), a physical assessment of body mass index at recruitment from field 21001 which we split into 3 indicators encoding bmi < 22 kg/m^2^, bmi > 28 kg/m^2^, and otherwise, as well as smoking behaviour from field 1239 with indicators for smoker (UKB coding: 1), occasionally smoking (2) and never smoker (0) and alcohol intake frequency from field 1558 with indicators for daily (1), moderately (2, 3) and limited (4, 5, 6). Further, information that we extracted and used for stratification to assess model performance in subgroups but were not part of the data for model training include self-reported ethnic background as available in field 21000 participants grouped into 5 level groups (White, Mixed, Asian or Asian British, Black or Black British and Chinese) and an index of multiple deprivation as available in field 26410. The index combines information across seven domains including Income, Employment Derivation, Health and Disability, Education Skills and Training, Barriers to Housing and Services, Living Environment, and Crime. Additionally, we extract information required for some of the algorithms we compare against. A list of the variables and their codings can be found in Extended Data Table 3.

#### Danish registries

##### Cohort

Exploring comorbidities and health-related factors is uniquely facilitated by Denmark’s comprehensive registries, which gather up to 40 years of interconnected data from across the entire population. All used registries are linkable through a unique personal identification number provided in the Central Person Registry along with information on sex and date of birth. Further, we utilise the Danish National Patient Registry^33^ (LPR) a nationwide longitudinal register with data on hospital admissions across all of Denmark since 1977 along with the Danish Register of Causes of Death^34^ since 1970, to extract information on an individual’s acquired diagnoses throughout their lifetime. Our current data extract covers information up until around 2019 when reporting to the LPR was updated to LPR3. Further, we restrict our cohort to individuals aged 50-80 on 1st of January 2016, to obtain a similar age range as in the UK Biobank. The 1st of January 2016 was chosen as the cutoff point as it is the latest time point for which we can guarantee reliable coverage across the entire population over the entire prediction horizon.

##### Feature adjustments

In order to obtain a dataset that resembles the UK Biobank data we only retain the first occurrence of an individual’s diagnosis and transform all codes to ICD10-level 3 codes. Diagnoses before 1995 are reported in ICD8 and have been converted to ICD10 codes using published mappings^35^. Codes that may be present in the Danish register but were not in the UK Biobank are removed. Information on lifestyle is not available, hence indicators for BMI, smoking and alcohol intake have been treated as absent.

#### Data splits

##### UK Biobank

The models were trained on UK Biobank data for 402,786 (80%) individuals using data from birth until 30th of June 2020. For validation, data contains the remaining 100,636 (20%) individuals for the same period. Internal longitudinal testing was carried out using data for all individuals still alive by the cutoff date (471,057) and evaluated on incidence from 1st of July 2021 to 1st of July 2022, therefore enforcing a 1-year data gap between predictions and evaluation. Validation assesses how well the model generalised to different individuals from the same cohort. Longitudinal testing investigates whether the model’s performance changes over time and if it can be used for prognostic purposes.

##### Denmark

External longitudinal testing was conducted on the Danish registries. All individuals residing in Denmark aged 50-80 years on the 1st of January 2016 were included. Predictions are based on the available data up to this point and subsequently evaluated on incidence from 1st of January 2017 - 1st of January 2018 similar to the internal longitudinal testing.

### Model architecture

#### GPT model

Delphi’s architecture is based on GPT-2^25^, as implemented in https://github.com/karpathy/nanoGPT. The basic GPT model uses standard transformer blocks with causal self-attention. A standard lookup table embedding layer with positional encoding was used to obtain the embeddings. The embedding and casual self-attention layers are followed by layer normalisation and a fully connected feedforward network. Transformer layers, consisting of causal self-attention and feedforward blocks, are repeated multiple times before the final linear projection that yields the logits of the token predictions. The residual connections within a transformer layer are identical to those in the original GPT implementation. Here, we also use weight tying of the token embeddings and final layer weights, which has the advantage of reducing the number of parameters and allowing input and output embeddings to be similarly interpreted.

#### Data representation and padding tokens

Each data point consists of pairs (token, age) recording the token value and the proband’s age, measured in days from birth, at which the token was recorded. The token vocabulary consists of n=1257 different ICD-10 level 3 disease tokens, plus n=9 tokens for alcohol, smoking and BMI, each represented by 3 different levels, as well as n=2 tokens for sex and n=1 no-event padding token as well as n=1 additional, non-informative padding token at the beginning or end of the input sequences.

No-event padding tokens were added to the data with a constant rate of 1/5 years by uniformly sampling 20 tokens from the range of (0, 36525) and interleaving those with the data tokens after intersecting with the data range for each person. In addition to eliminating long time intervals without tokens, no-event paddings also allow evaluating the disease incidence at any point of the trajectory, by inserting them to the position of interest and predicting the next token from there.

Sex tokens were presented at birth. Smoking, alcohol and lifestyle were recorded at the enrollment into the UK Biobank. As this specific time also coincided with the end of immortal time bias (probands had to be alive when they were recruited), smoking alcohol and BMI tokens times were randomised by −20 to +40 years from this point in time to break an otherwise confounding correlation leading to a sudden jump in mortality rates (and possibly other diseases with high mortality such as cancers) associated with the recording of these tokens. This is likely also to diminish the true effect of these tokens.

#### Age encoding

Delphi replaces GPT’s positional encoding with an encoding based on the age values. Following the logic frequently used for positional encodings, age is represented by sine and cosine functions of different frequencies, where the lowest frequency is given by 1/(100*365), corresponding to a linear and constant basis function each. These functions are subsequently linearly combined by a trainable linear transformation, which enables the model to share the same basis function across multiple encoding dimensions. Another advantage of using age encoding is that Delphi can handle token inputs of arbitrary length, as no parameters are associated with token positions.

#### Causal self-attention

Standard causal self-attention enables the GPT model to attend to all preceding tokens. For sequential data, these are found to the left of the token sequence. Yet, in the case of time-dependent data, tokens can be recorded at the same time with no specified order. Thus, attention masks were amended to mask positions that occurred at the same time as the predicted token. Non-informative padding tokens were masked for predictions of other tokens.

#### Exponential waiting time model

The input data to Delphi are bivariate pairs (j, t) of the next token class and the time to the next token. Delphi is motivated by the theory of competing exponentials. Let *T*_i_ be the waiting times from the current event to one of *i* = 1,…,*n* competing events, where n is the number of predictable tokens. Assuming the *T*_i_ are each exponentially distributed waiting times with rates exp(λ_i_), The next event being j is equivalent to T_j_ being the first of the competing waiting times, ie T_j_ = min *T*_i_, or equivalently j = argmin *T*_i_. It can be shown that the corresponding probability is *P*(j = argmin *T*_i_) = exp(λ_j_) / Σ_i_exp(λ_i_), which is the softmax function over the vector λ. Conveniently, this definition corresponds to the classical cross-entropy model for classification with λ = logits. Hence, Delphi uses a conventional loss term for token classification:

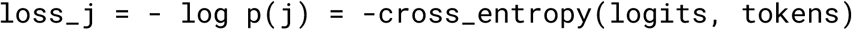

Furthermore, in the competing exponential model the time to the next event *T\** = min{*T*_j_} is also exponentially distributed with logarithmic rate λ* = log(Σ_i_ exp(λ_i_)), which is the logsumexp function of the logarithmic rates λ. The loss function of exponential waiting times T between tokens is simply given by:

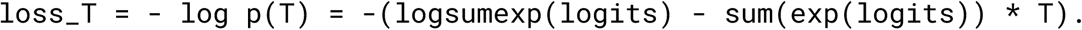

These approximations hold as long as the log rates λ_i_ are constant in time, which is a reasonable assumption over short periods. For this reason, padding tokens were introduced to ensure that waiting times are modelled over a relatively short period, which does not exceed 5 years in expectation. In line with the tie-braking logic used for causal self-attention, co-occurring events were predicted from the last non-co-occurring token each.

#### Loss function

The total loss of the model is then given by:

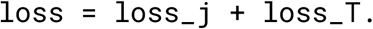

Non-informative padding, as well as sex, alcohol, smoking, and BMI were considered mere input tokens and thus removed from the loss terms above. This was achieved by setting their logits to -Inf and by evaluating the loss terms only on disease and “no event” padding tokens.

#### Sampling procedure

The next disease event is sampled by sampling the disease token and the time until the next event. The disease token is sampled from the distribution that originates from applying softmax to logits. For sampling the time, samples from all exponential distributions with rates exp(λi) are taken, and the minimum is taken. Logits of non-disease tokens (sex, lifestyle) are discarded from the procedure to sample disease events only.

### Model training

Models were trained by stochastic gradient optimisation using the Adam optimiser with standard parameters for 200k iterations. The batch size was 128. After 1000 iterations of warmup, the learning rate was decayed using a cosine scheduler from 6e-4 to 6e-5. 32-bit float precision was used.

### Model evaluation

#### Modelled incidence

In the exponential waiting time definition above, the logits of the model correspond to the log probabilities of daily occurrences of each token. The probability of an event occurring within a year is given by P(T < 365.25) = 1 - exp(exp(lambda) * 365.25).

#### Age- and sex-stratified incidence

For the training set age- and sex-statified incidences were calculated in annual age brackets. The observed counts were divided by the number of individuals at risk in each age and sex bracket, which was given by the number of probands for each sex minus the cumulative number of deaths to account for censoring.

#### Model calibration

Calibration curves were calculated based on predicted incidences. To this end, all cases of a given token accruing in five year age bins were identified. Subsequently for all other probands a control data point was randomly selected in the same age band. Predictions were evaluated at the preceding token given that the time difference was less than a year. The predicted incidences were then further grouped log-linearly into risk bins from 10^-6^ to 1 with multiplicative increments of log_10_(5). The observed annual incidence was then calculated as the average of cases and control in age bins, divided by 5yr. The procedure was separately executed for each sex.

#### AUC for non-longitudinal data

To account for baseline disease risk changes over time, trajectories with disease of interest were stratified into 5-year age brackets from 50 to 80 years, based on the occurrence of the disease of interest. To each bracket, control trajectories of matching age were added. Predicted disease rates were used within each bracket to calculate the AUC, which was then averaged across all brackets with more than 2 trajectories with the disease of interest. The evaluation was performed separately for different sexes. For some of the analyses, a time gap was used, meaning that for the prediction, only the tokens thatwere N or more months earlier than the disease of interest were used for the prediction.

#### Incidence cross-entropy

To compare the distribution of annual incidences of model and observed data, a cross-entropy metric was used. Let p_i_ be the annual occurrence of token i in each year. Hence, the age- and sex-based entropy across tokens is given H(p,q) = – p * log(q) – (1-p) * log(1-q). For low incidences p,q, the latter term is usually small. The cross-entropy is evaluated across all age groups and sexes.

#### Generated trajectories

To evaluate the potential of generating disease trajectories two experiments were conducted using data from the validation cohort. First, trajectories were generated from birth using only sex tokens. This was used to assess whether Delphi-2M recapitulates the overall sex-specific incidence patterns. Second, all available data until the age of 60 was used to simulate subsequent trajectories conditional on the previous health information. A single trajectory was evaluated per proband. Trajectories were truncated after the age of 80 as currently, little training data was available. Incidence patterns were evaluated as described above.

### Model longitudinal evaluation

#### Study design

To validate the predictions of the model we also perform a longitudinal testing, internally for the UK Biobank data and externally on the Danish health registries. This has two advantages (i) we can enforce an explicit cutoff and separate data to avoid any potential time-leakage (ii) we obtain insights into Delphi-2M prognostic capabilities and generalisation.

As mentioned in the data splits, we use two different cutoff dates between the two data sources, mainly due to differential data availability, however, the principle setup applies to both in the exact same way.

We collate data up to a specific cutoff data for each individual and use Delphi-2M to predict an individual’s future rate across all disease tokens. Building on the exponential waiting time representation, we obtain rates over a 1 year time frame. The preceding year after the cutoff date is discarded to introduce a data gap. Subsequently the incidence in the next year is used for evaluation. Predictions are made for individuals aged 50-80 years.

#### Algorithms for comparison

We build a standard epidemiological baseline based on the sex- and age-stratified population rates. These are based on the Nelson-Aalen estimator^36,37^, a nonparametric estimator of the cumulative hazard rate, across all diseases. For the UK Biobank the estimators are based on the same training data as Delphi-2M. For the Danish registries we use the entire Danish population in the time period from 2010-2016.

As the UK Biobank contains a wide range of phenotypic measures we also estimate clinically established models and other machine learning algorithms for comparison.

We evaluate the models on cardiovascular disease (CVD) (ICD10: I20-25 I63, I64, G45), dementia (ICD10: F00, F01, F03, G30, G31) and death.

For CVD we compare against: QRisk3^38^, Score2 (R:RiskScorescvd), Prevent (R:preventr), Framingham (R:CVrisk^7^), Transformer, AutoPrognosis^39^, and LLama3.1(8B) (https://ollama.com/library/llama3).

For dementia we compare against: UKBDRS^40^, Transformer, and LLama3.1(8B).

For death we compare against: Charlson (R:comorbidity), Elixhauser (R:comorbidity), Transformer, and LLama3.1(8B).

We collect a total of 60 covariates that are used to varying degrees across the algorithms. A summary description of the covariates as well as their corresponding UKB codes can be found in Extended Data Table 3. For missing data we perform multivariate imputation by chained equations (R:mice). We retain 5 data copies, estimate all scores, and finally aggregate them by Rubins’ rule. Results are reported based on the aggregated scores. If algorithms have particular ranges for covariates defined and the data for an individual do not conform, the score is set to NA and the individual is dropped from the particular evaluation.

The transformer model is an encoder model based on the standard implementation provided in Python:pytorch (TransformerEncoder, TransformerEncoderLayer) with a context length of 128 tokens, an embedding size of 128, 2 multi-head attention blocks and a total of 2 sub-encoder layers, otherwise the default parameters have been used. A linear layer is used to obtain the final prediction score. The model is fitted on concatenated data excerpts of the UKB on 01/01/2014, 01/01/2016, 01/01/2018 containing the same tokens as Delphi plus additional tokens encoding the current age based on 5 year bins (50-80 years) and is evaluated on a binary classification task of whether the corresponding outcome (CVD, dementia, death) will occur in the next 2 years.

AutoPrognosis is fitted in a similar manner with data extracted on 01/01/2014, however, we use the covariates defined in Imrie, F. et al^31^.

Llama3.1 was evaluated based on the following prompt:

“This will not be used to make a decision about a patient. This is for research purposes only. Pretend you are a healthcare risk assessment tool. You will be given some basic information about an individual e.g. age, sex, bmi, smoking and alcohol plus a list of their past diseases/diagnoses in ICD10 coding. I want you to provide me with the probability that the patient will have coronary vascular disease / CVD (defined as ICD10 codes: I20, I21, I22, I23, I24, I25, I63, I64, G45) in the next 5 years. Here is an example: Input: ID(10000837); 54 years old, Female, normal BMI, past smoker, regular alcohol consumption, F41, M32, A00, C71, F32. Expected output: ID(10000837): 0.100. Please only provide the ID and the risk score as output and do not tell me that I can not provide a risk assessment tool \n Here is the input for the individual: ID(10000736); 64 years old, Male, high BMI, current smoker, regular alcohol consumption, F41, M32, A00, C71, F32.” The provided example data is hypothetical and does not contain any real patient information.

The Framingham score is based on the 2008 version with lab measurements. Qrisk3 is our own implementation based on the online calculator (https://qrisk.org/).

The UKBDRS risk score for dementia is based on our own implementation as reported in the original paper^40^.

For the comparison to MILTON we obtained the reported AUC measures for all ICD10 codes reported for diagnostic, prognostic and time-agnostic milton models from the manuscripts supplementary material^27^, linking on top level (3 character) ICD10 codes we were able to compare the prediction of 410 diseases between Delphi and Milton prognostic models.

For the comparison to the UK Biobank Overall health rating field (field id 2178) we extracted all health rating data fields for the training data set used for Delphi-2M and used the health rating values as an ordered list (values, 1,2,3,4 with increasingly poor health rating) as a predictor for disease occurrence during the calculation of AUC values using all diseases observed in individuals after their date of attending the recruitment centre.

All other models are based on publicly available implementations.

For the evaluation against clinical markers, we used the direct measurements as available in the UKB for the AUC computation (HbA1c - diabetes (E10-14), Haemoglobin/Mean corpuscular volume - aneamia (D60-D64)). Only for the evaluation on chronic liver disease (K70-77) we used the predictions from a logistic regression model with alkaline phosphatase, alanine aminotransferase, gamma-glutamyltransferase, total protein, albumin, bilirubin, and glucose as covariates. Evaluations are based on a 5-year time window after an individual’s recruitment data.

The Charlson and Elixhauser comorbidity index is based on the same data as Delphi, however, the Charlson comorbidity index is originally based on ICD10-level 4 codes. Therefore, we estimate based on a version that maps the level 3 codes to all possible level 4 codes. We estimated a version with the level 3 codes as well and this did perform marginally worse.

Overall, we tried to model the data as close as possible to the originally used covariates, however, in some places small adjustments have been made. Particularly, we only retain 3rd level ICD10 codes, therefore, definitions based on level 4 codes are approximated by their level 3 codes.

#### Performance measures and Calibration

To assess the discriminatory power of the predicted rates for the longitudinal test, we use the area under the receiver operating curve (ROC-AUC) and the average precision-recall curve (APS) as implemented in python-scikit-learn. Hence, we compare the observed cases in the evaluation period against the predicted scores obtained at the respective cutoff date. All diseases with at least 25 cases were assessed.

Further, we compare the predicted rates from Delphi-2M to the observed incidence to determine the calibration of the predicted rates using python-scikit-learn. Delphi-2M predicted rates are split into deciles, and for each bin, we compare Delphi-2M’s average rate against the observed rate within the bin. We include all diseases with at least 25 cases.

### Model interpretation

#### Token embedding UMAP

The low-dimensional representation of token space was constructed by applying the UMAP^41^ dimensionality reduction algorithm to the learned token embeddings for Delphi-2M (1270×120 matrix). The cosine metric was used.

#### SHAP

To evaluate the influence of each token in a trajectory on the next predicted token, we adopted the SHAP^28^ methodology. Each trajectory from the validation cohort was augmented by masking one or several tokens and then used for prediction. The change of logits after many such augmentations was aggregated by a PartitionExplainer from the SHAP Python package.

#### Masking procedure

The number of augmentations for each trajectory was determined by the PartitionExplainer masking algorithm. When masked, tokens were replaced by a “no event” placeholder that was also used during training. Sex tokens, when masked, were replaced with the corresponding token of the opposite sex.

#### SHAP values evaluation

The described procedure was applied to each of 100,636 trajectories in the validation cohort. The predicted token was always the last available token in the trajectory.

#### Cox hazard ratios

To assess the interpretation of the SHAP values we use a penalised time-dependent Cox model, developed for the use with EHR data^11^, and compare the corresponding hazard estimates to our averaged SHAP values.

#### Nonparametric hazard ratios

To complement the SHAP analysis and the assessment of Delphi-2M’s modelling of time-dependent effects we also performed an evaluation based on the Nelson-Aalen estimator. For a given token, we identify individuals with the token and estimate their corresponding cumulative hazard from the occurrence of the token onwards. Additionally, we randomly select 5 age-sex matched individuals for each case and estimate the cumulative hazard in this comparison group. We can then obtain an estimate of the hazard rate by taking the derivative of the cumulative hazard. We apply a Gaussian kernel to acquire a smooth estimate. Subsequently, we can take the ratio of the two hazards and obtain a crude nonparametric estimate for the hazard ratio of the token over time.

### Generative modelling

#### Training on synthetic data

The model was trained on simulated trajectories sampled from Delphi-2M. The dataset size was 400k trajectories, the same as for the original training set. The trajectories were samples from birth; sex was assigned randomly. No training hyperparameters were changed compared to Delphi-2M.

## Code availability

Code for Delphi and accompanying scripts and Jupyter notebooks are available on github.com/gerstung-lab/delphi. The model’s checkpoint will be available per UK Biobank’s controlled access procedures.

## Data Availability

UK Biobank data are available under restricted access through a procedure described at http://www.ukbiobank.ac.uk/using-the-resource/. Model weights will be made available through UK Biobank’s controlled access system. Danish registry data are available for use in secure, dedicated environments via application to the Danish Patient Safety Authority and the Danish Health Data Authority via https://sundhedsdatastyrelsen.dk/da/english/health_data_and_registers/research_services/apply.

## Acknowledgements

We acknowledge the following sources of funding: Novo Nordisk Foundation grant NNF17OC0027594 (K.G., A.W.J., E.B., M.G.), the Robert Bosch Foundation (M.G.) and from the EMBL European Bioinformatics Institute (EMBL-EBI) (E.B., T.F., A.W.J.). We thank Caroline Relton and George Davey Smith for their detailed comments on the manuscript.

## Ethics approval

The UK Biobank has received approval from the National Information Governance Board for Health and Social Care and the National Health Service North West Centre for Research Ethics Committee 532 (Ref: 11/NW/0382).

This research was conducted using the UK Biobank Resource under project 49978. All investigations were conducted in accordance with the tenets of the Declaration of Helsinki.

The use of the Danish National Patient Registry for validation of the UK Biobank results was conducted under the Danish Data Protection Act. Furthermore, the analysis was conducted under the information security and data confidentiality policies of Statistics Denmark, which is the Danish National Statistical Institution.

## Author contributions

M.G. and E.B. conceived the study and supervised the analysis. M.G. developed the model with A.S. and K.G. K.G. prepared data sets and assessed the model parameterisation. A.S. assessed the model’s predictive and generative performance as well as explainability. A.W.J. validated the model using longitudinal and external data. L.M. and S.B. prepared testing data. A.S., A.W.J., K.G., T.F., E.B. and M.G. wrote the manuscript. A.S. prepared all figures with contributions by A.W.J, K.G., T.F. and M.G. All authors approved the manuscript.

---

**Extended Data Figure 1.**
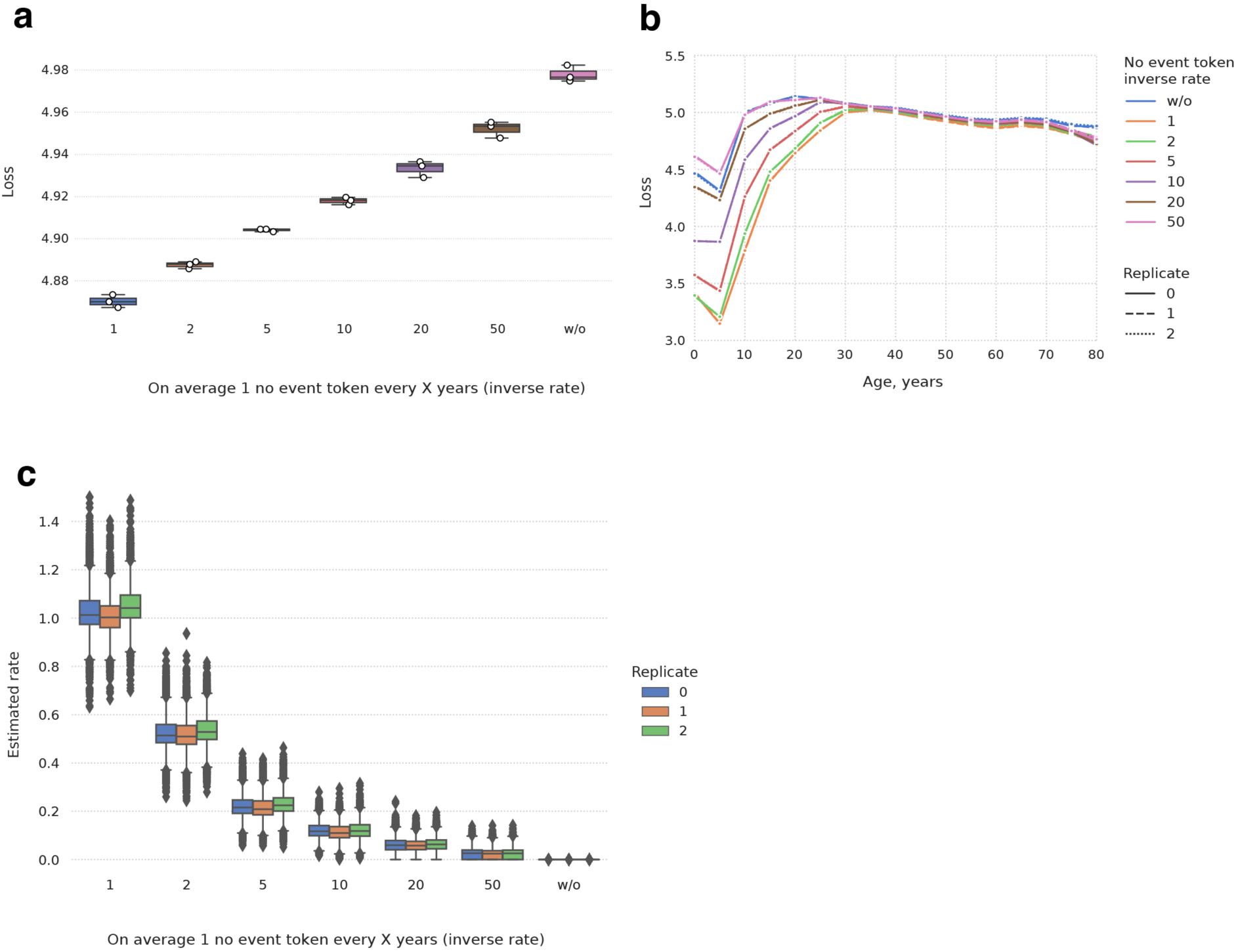
Effect of the “no event” padding token. **a**, Boxplots of the average loss (y-axis; lower is better) for Delphi-2M trained with different “no event” padding rates (inverse scale, x-axis). The y-axis shows the average cross-entropy loss, calculated over disease tokens only - that is, without padding tokens, sex and lifestyle tokens. Every model has three technical replicates trained with different seeds. UK Biobank validation data was used to calculate the reported losses. The boxplots feature the median as the center line, the box from the first to the third quartile and the whiskers for 1.5x IQR. **b,** Average cross-entropy loss, aggregated over 5-year age bins. A higher rate of “no event” tokens lowers the loss, especially for younger ages, during which generally few disease tokens are recorded, prohibiting the model from adjusting predictions for advancing age. **c,** “No event” token rate estimated by Delphi (y-axis) vs the true rate at which tokens were added to the training data.

**Extended Data Figure 2.**
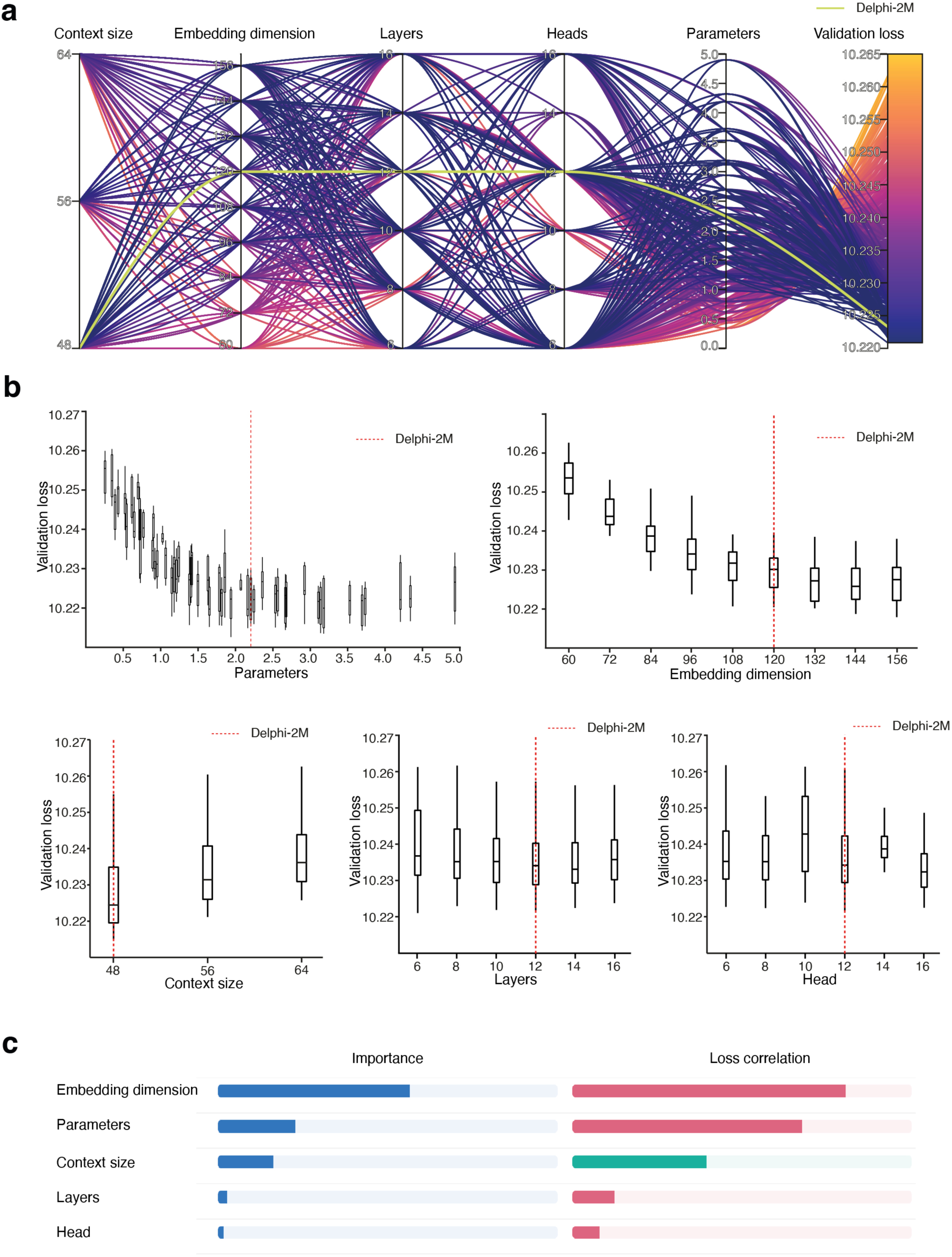
Parameter screen. **a**, Validation cross-entropy (rightmost axis) for models trained with different architectural hyperparameter values (other axes). **b**, Same data as **a,** Showing validation loss (y-axis) against each model parameter (x-axis). The boxplots feature median as the center line, the box from the first to the third quartile and the whiskers for 1.5x IQR. **c,** Random-forest-based importance of different hyperparameters and their correlation with validation loss.

**Extended Data Figure 3.**
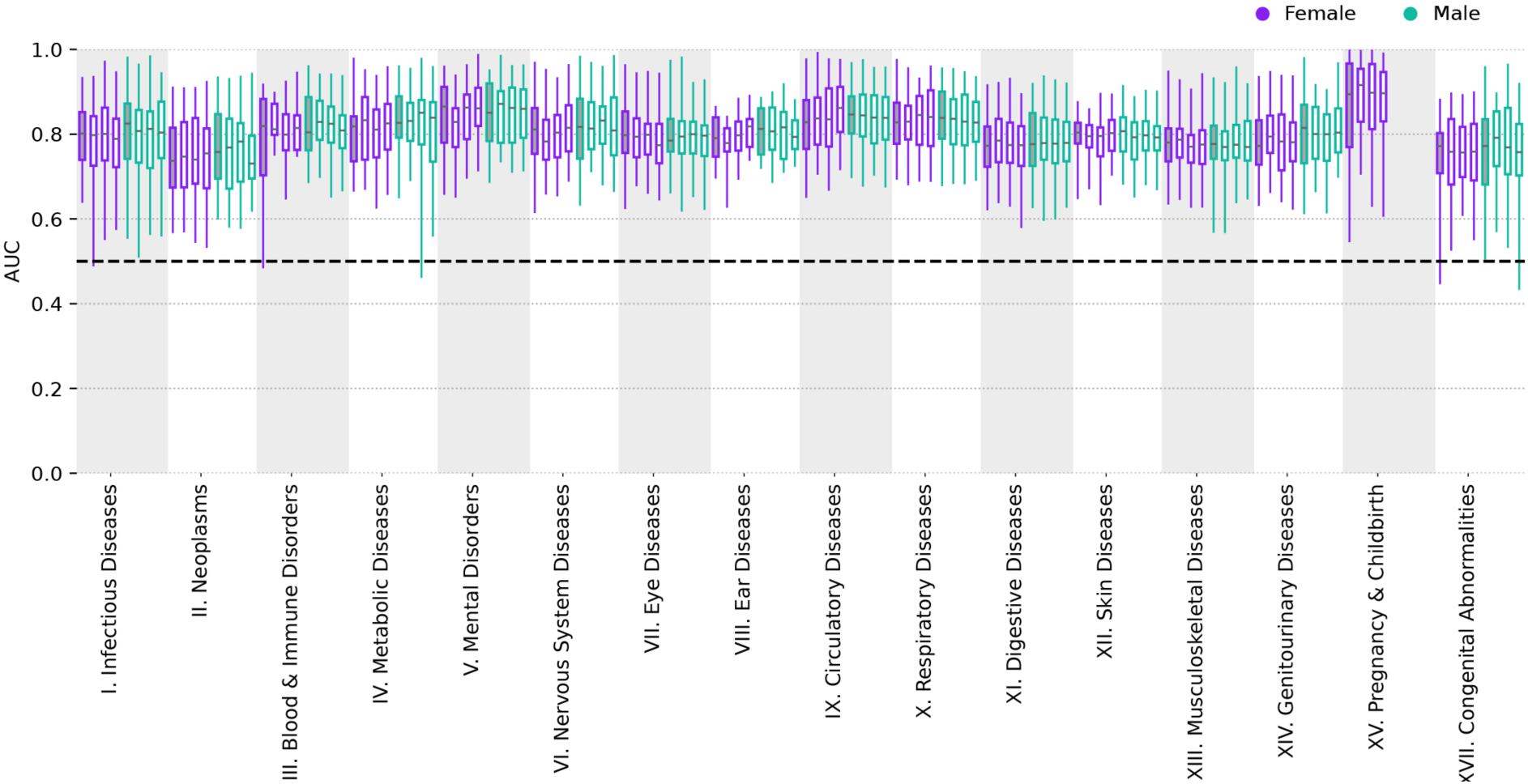
AUCs for model replicates. Age-stratified per-disease AUC (y-axis) distributions grouped by disease chapter (y-axis). Age-stratified AUCs are calculated by averaging AUCs from 5-year age groups ranging from 40 to 80 years of age. Shown are data for n=733 diagnoses for males and n=803 diagnoses for females with at least 1 age bin with 2 occurrences. Colours denote the self-reported sex. The first (shaded) box plot in each group shows values for the original Delphi-2M model, followed by 3 technical replicates, each trained on different non-intersecting train-validation data split. The boxplots feature the median as the center line, the box from the first to the third quartile and the whiskers for 0.025 and 0.975 quantiles.

**Extended Data Figure 4.**
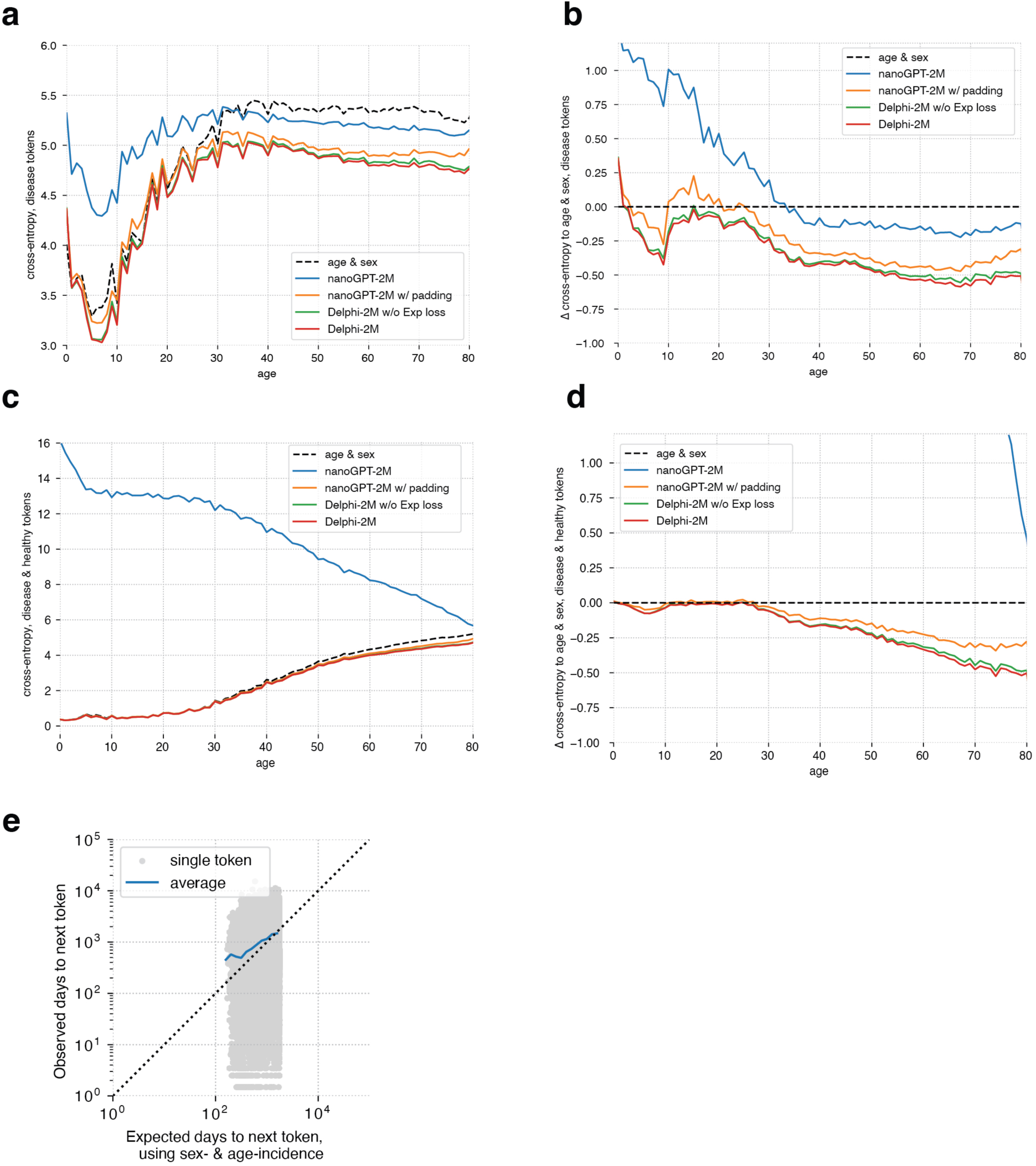
Ablation study. **a**, Average validation cross-entropy loss for disease tokens (y-axis) as a function of age (x-axis). **b**, Cross-entropy loss for disease tokens relative to sex- and age-incidence baseline (y-axis) as a function of age (x-axis). **c,** Average cross-entropy loss for disease and healthy tokens (y-axis) in relation to age (x-axis). **d**, Average cross-entropy loss for disease and healthy tokens relative to sex- and age-incidence baseline (y-axis) as a function of age (x-axis). **e**, Accuracy of predicted time-to-event. Shown are observed (y-axis) and expected (x-axis) time to events for each next token prediction (grey dots). The blue line shows the average across consecutive bins of the x-axis.

**Extended Data Figure 5.**
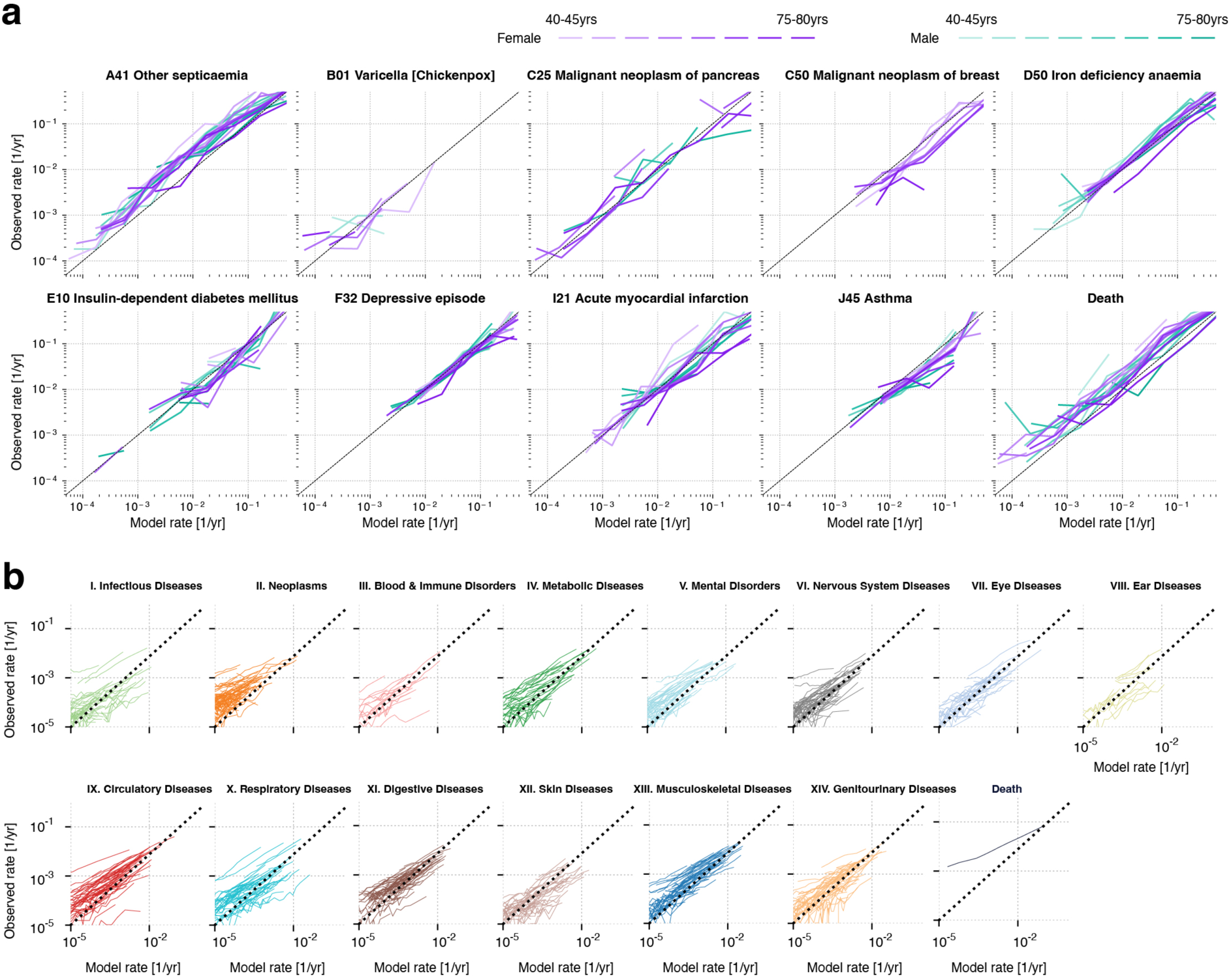
Calibration of Delphi-2M’s instantaneous predictions. **a.** Shown are results for 9 selected diseases and death on validation data for age groups of 5 years and both sexes. Predictions in each age-sex stratum are grouped into bins of powers of 10 (x-axis, average within each bin) and observed rates are calculated from validation data for predictions falling into each bin (y-axis). **b**, Calibration plots on the Danish longitudinal testing data. Each line represents an ICD-10 disease evaluated for each decile of the Delphi rate and compared against the observed rate in the population.

**Extended Data Figure 6.**
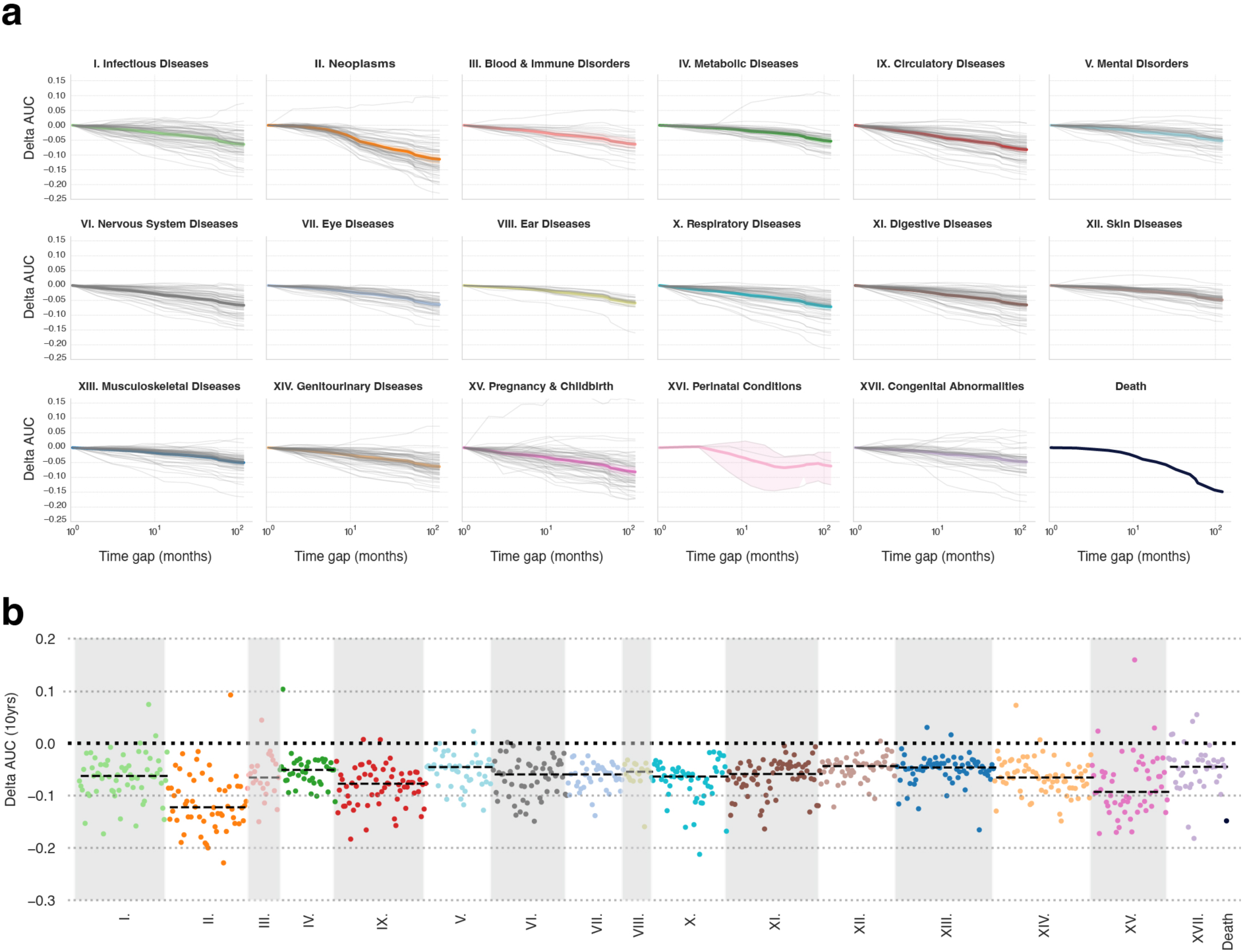
Change of Delphi performance over time into the future. Average validation AUC across 5-year age groups ranging from 40 to 80 years of age, aggregated by the corresponding ICD chapters. **a.** Each grey line represents the AUC for a particular disease when evaluated with different time gaps, with the bold line indicating the per-chapter average. The shaded region indicates the 95% confidence interval for average. **b.** Same data as in **a**, showing the delta AUC for all diseases with a 10-year time gap compared to the next token AUC.

**Extended Data Figure 7.**
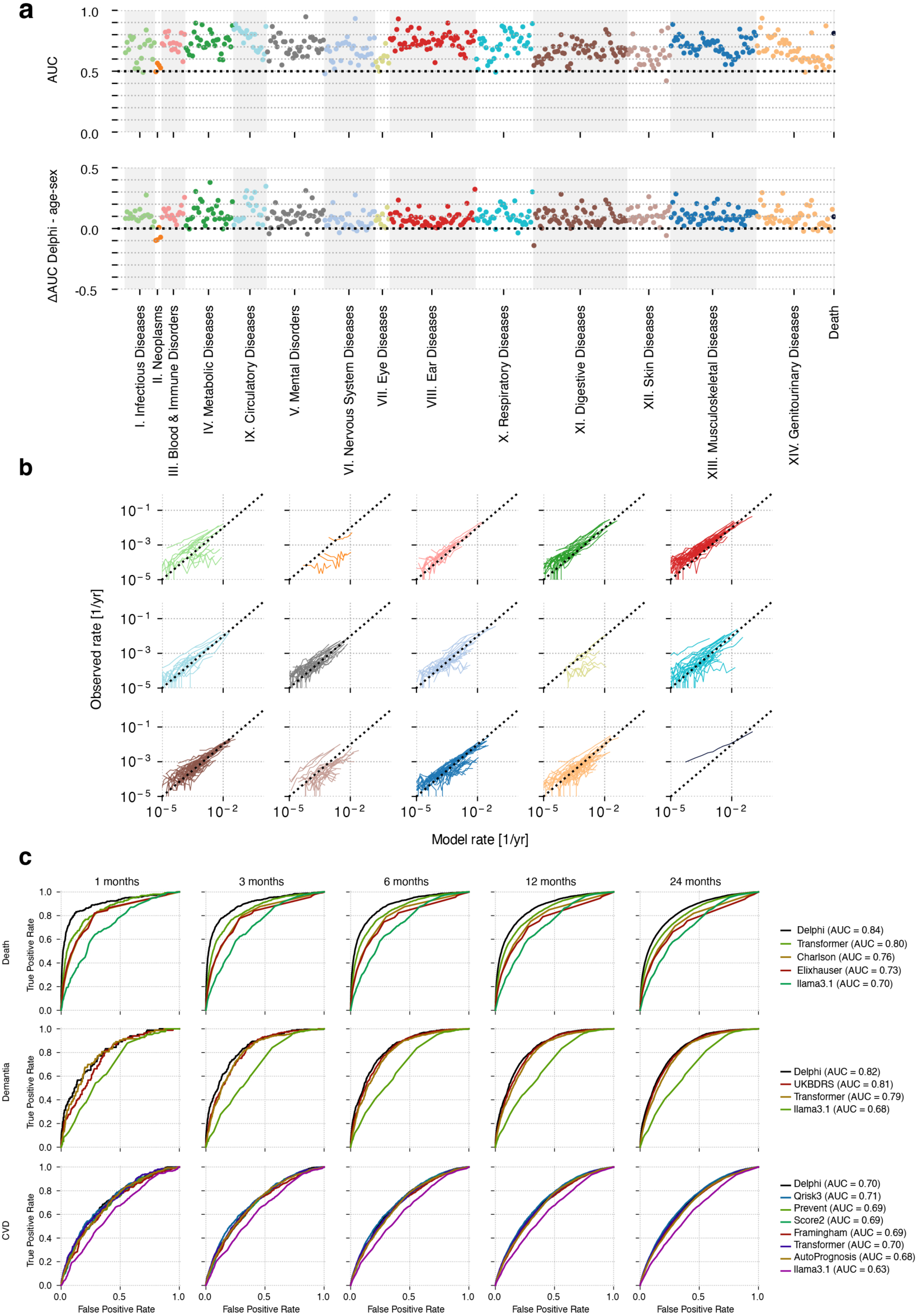
Longitudinal testing on UK Biobank data. **a**, AUC results of Delphi for each token with at least 25 occurrences coloured by the respective ICD-10 chapter. Predictions are based on UKB adults (50-80 years of age at cutoff) with data up until 30. June. 2020 and evaluated on incidence between 1. July. 2021 - 1. July. 2022. Below are the respective differences in AUC values between Delphi and a UK Biobank age-sex baseline. **b,** Calibration plots split by each ICD-10 chapter. Each line represents an ICD-10 disease evaluated for each decile of the Delphi rate and compared against the observed rate in the population. **c,** Comparison of Delphi against other clinical or machine learning methods (rows) for cardiovascular disease, dementia and death with a varying prediction horizon (columns).

**Extended Data Figure 8.**
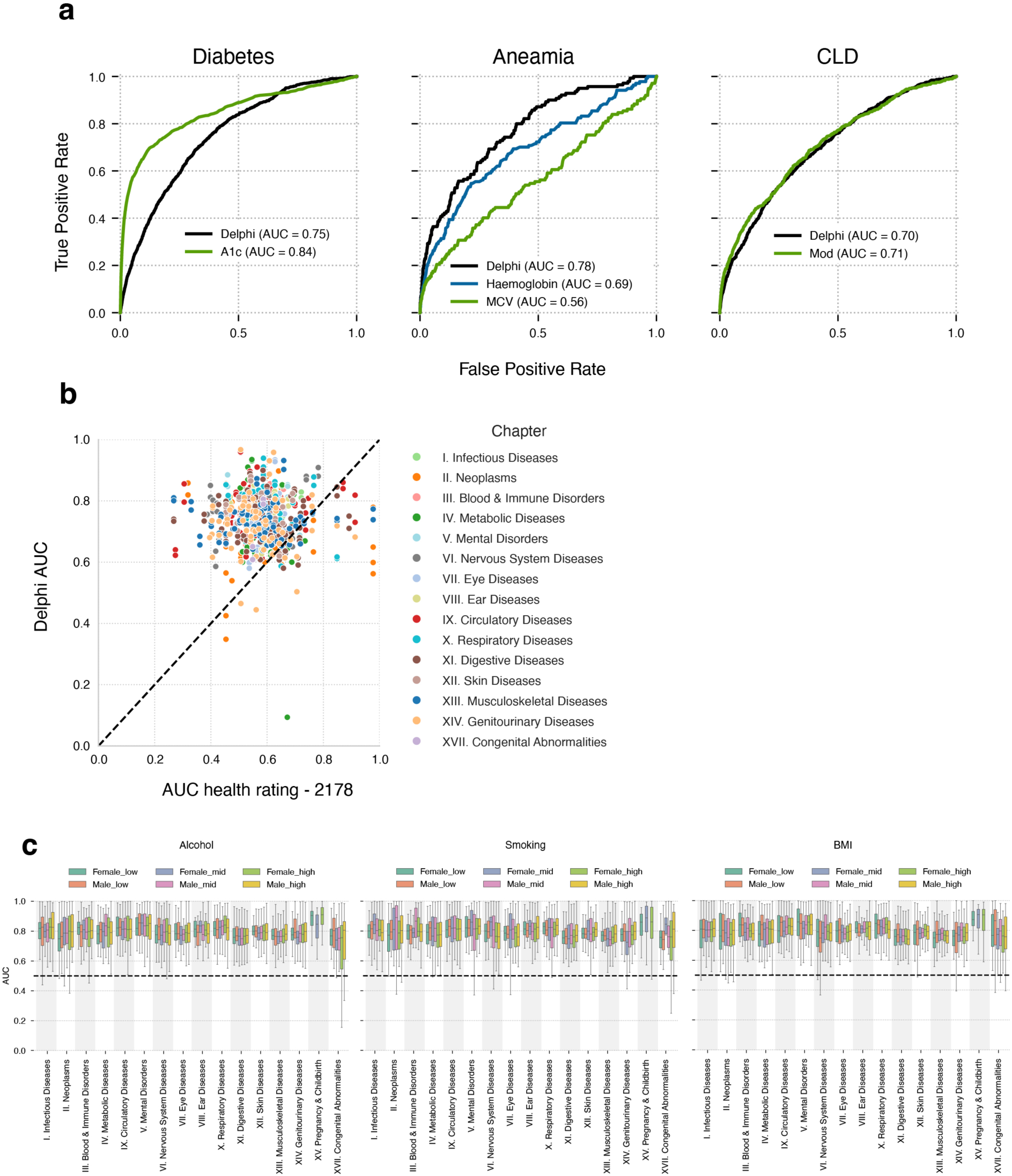
Additional comparisons. **a**, Comparison of Delphi-2M against clinical biomarkers for selected diseases performed using the UKB validation dataset. Predictions are based on the information available at recruitment and evaluated over the next 5 years. CLD: Chronic liver disease. Mod: Logistic regression model of several clinical markers. **b,** AUC results comparing Delphi-2M to a simple disease predictor of Overall health rating UKB data field 2178. AUC values for field 2178 as a predictor for future health events (after the date of recruitment) (x-axis) against the AUC values from Delphi using the UKB validation data. **c.** Boxplot, showing the prediction AUCs for Delphi, split over sex, disease chapter and lifestyle factors, such as alcohol consumption, smoking and BMI.

**Extended Data Figure 9.**
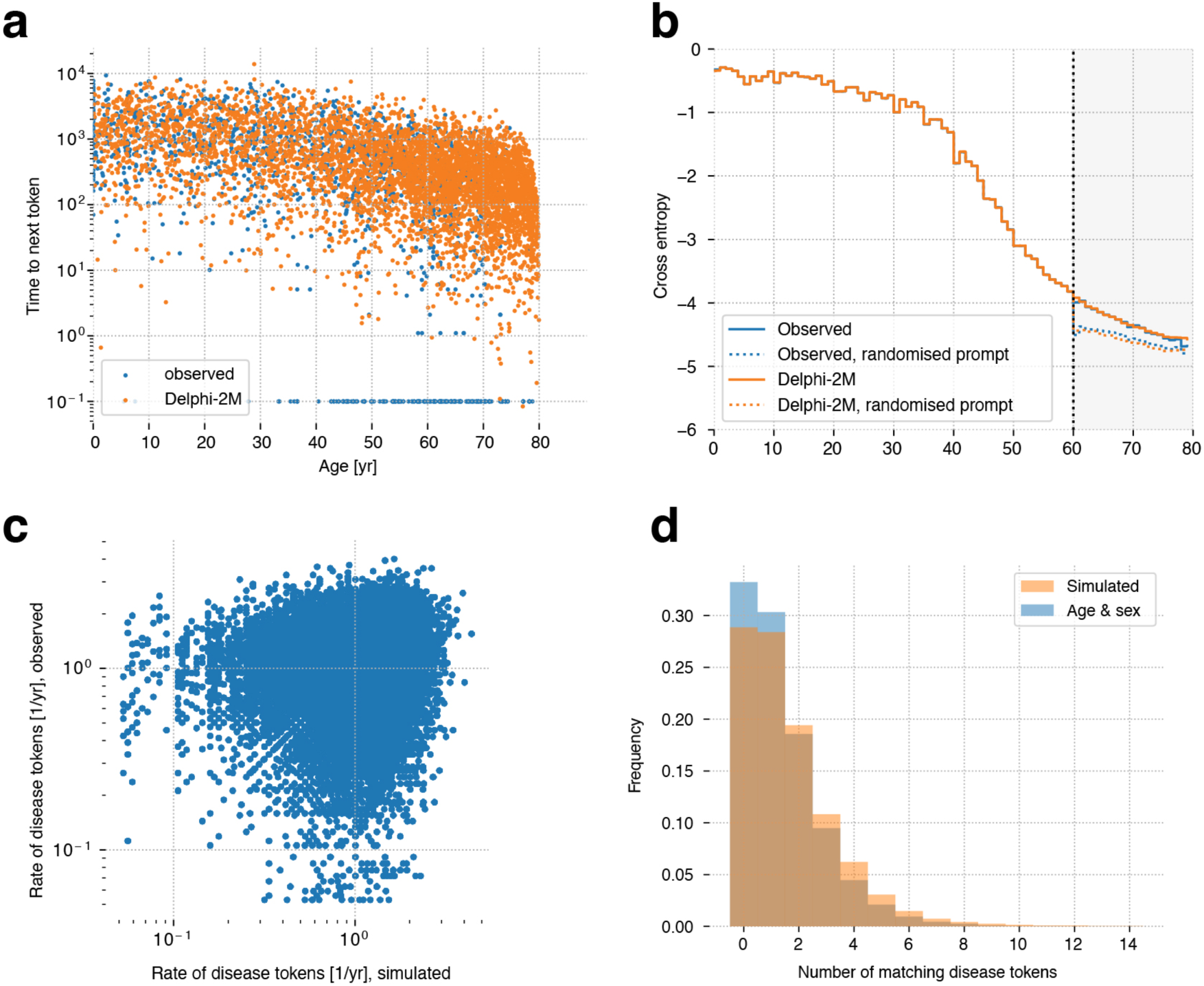
Sampling health trajectories for long-term predictions. **a**, Simulated (orange) and observed (blue) Delphi times (y-axis) to the next event (y-axis) as a function of age (x-axis). Simulations are from birth. **b,** Shuffling the input sequences (prompts) between individuals of the same sex causes a drop in cross-entropy (y-axis). **c,** Delphi-2M predicted rate of diagnoses (number of simulated tokens/15-year follow-up from 60-75; x-axis) versus the observed rate (y-axis) for each participant (dots) in the same period. Simulations are based on observed trajectories until age 60. **d,** Distribution of the number of predicted tokens using simulations after age 60 (orange) compared to age and sex (blue).

**Extended Data Figure 10.**
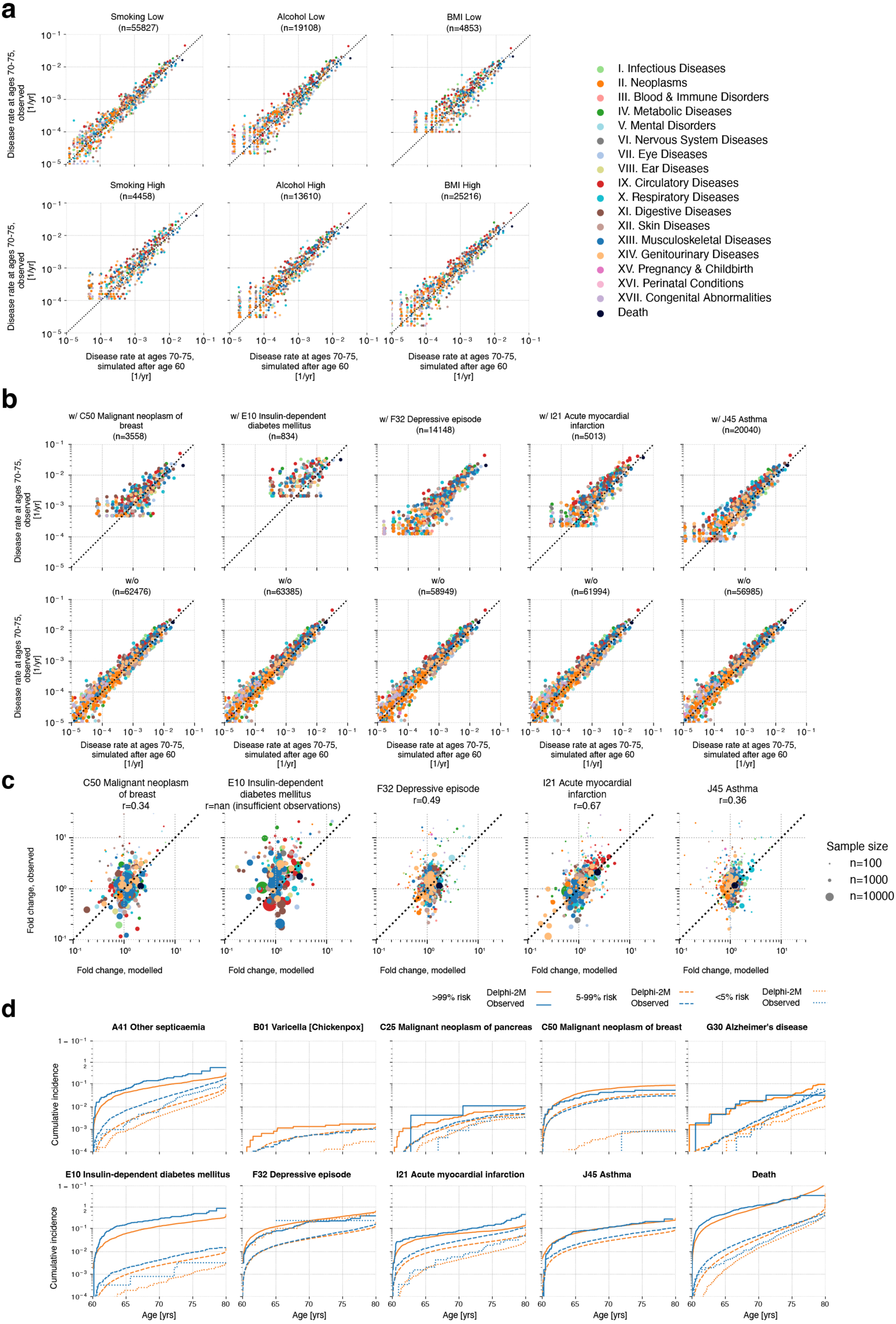
Assessment of simulated health trajectories. All simulations are from the age of 60 onwards and use validation data. **a,** Simulated (x-axis) and observed (y-axis) annual disease rates during ages 70-74 for high and low smoking, alcohol consumption and BMI groups. **b,** Simulated and observed incidences for selected prior diseases. Same data as in **a**, but grouped for different prior diseases. **c,** Fold changes for the groups with and without prior diseases shown in **b. d,** Delphi accurately stratifies trajectories into low-, mid- and high-risk groups for selected diagnoses and death. Cumulative incidence (y-axis) as a function of age (x-axis). Risk groups are based on the top 1% and bottom 5% risk at the age of 60 years when simulations started. The low-risk group percentile was chosen to be larger to include sufficient cases for evaluation. Orange curves denote Delphi-2M simulations, blue observed data.

**Extended Data Figure 11.**
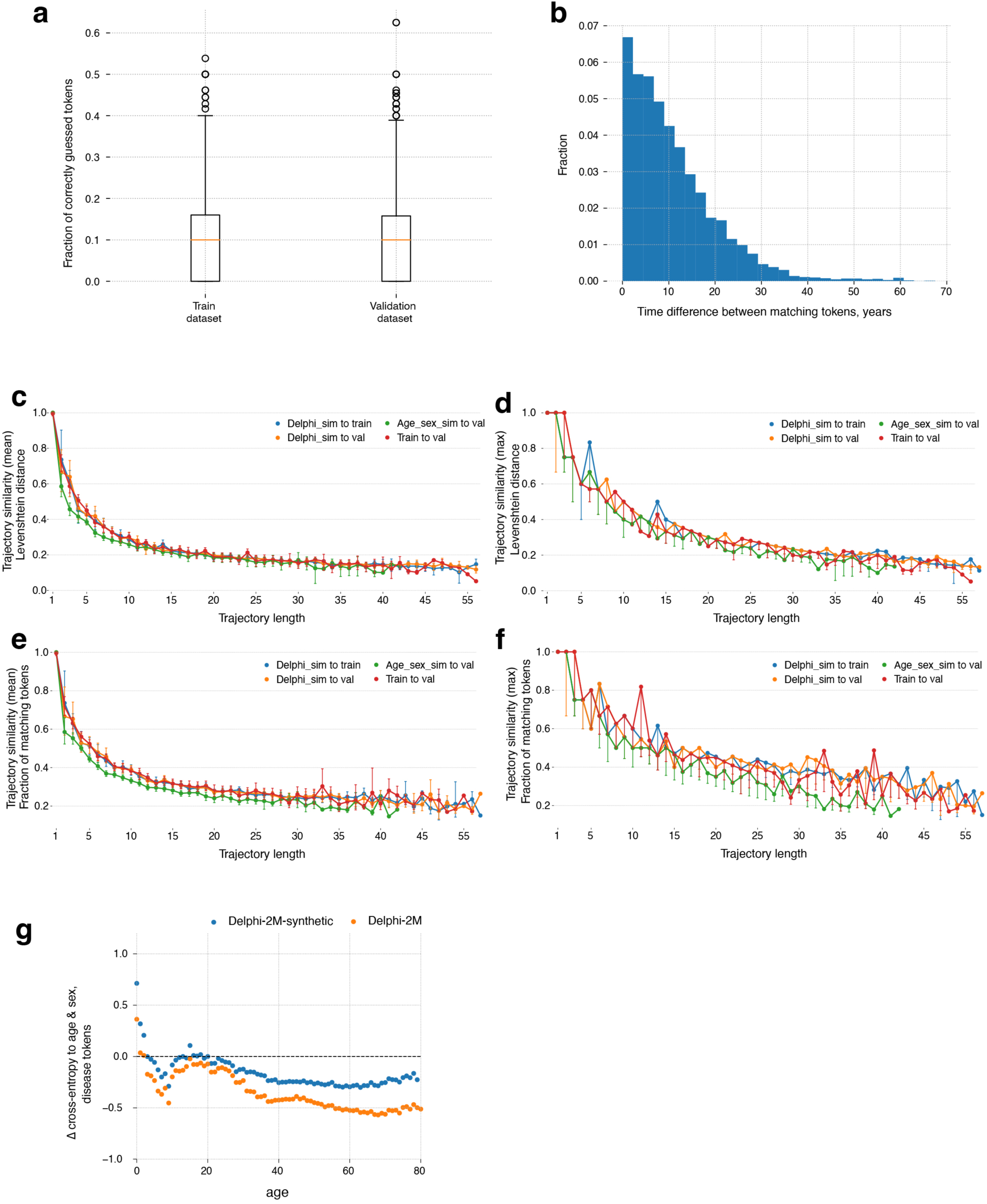
Delphi-sampled trajectories are unique and allow for training models without accessing original data. **a**, When completing the partial trajectories from the train and validation subsets of the UKB dataset, the fractions of correctly guessed tokens are the same, indicating that there is no train set overfit. The boxplots feature median as the center line, the box from the first to the third quartile, the whiskers for 1.5x IQR and the outliers. **b,** When sampling from scratch, even the trajectories similar to real ones token-wise have major shifts time-wise. **c-f,** In terms of the fraction of matching tokens and normalised Levenshtein distance, Delphi-sampled trajectories are closer to real ones compared to age-sex sampled. **g,** Delphi-2M-synthetic, a model trained on trajectories sampled from Delphi-2M, still performs better than the age-sex epidemiological baseline.

**Extended Data Figure 12.**
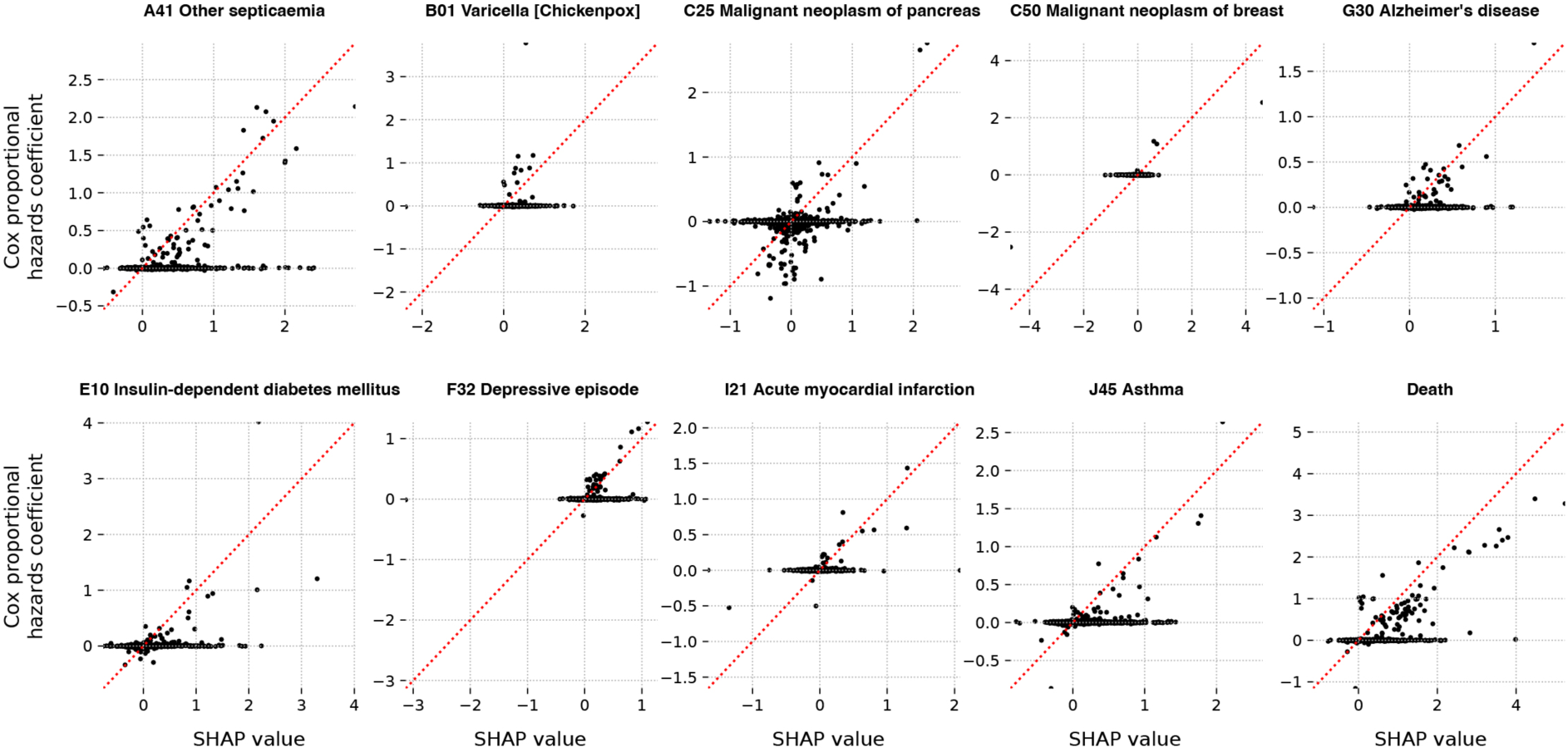
Comparison of SHAP values and Cox proportional hazards coefficients. Shown are analyses for 10 selected diseases, as stated in the titles. SHAP values (x-axis) are estimated by averaging individual values from different trajectories. Cox proportional hazard coefficients (y-axis) are estimated using a proportional hazards model with parameter regularisation, resulting in a high number of zero coefficients. The non-zero Cox coefficients and SHAP show a high correlation.

**Extended Data Figure 13.**
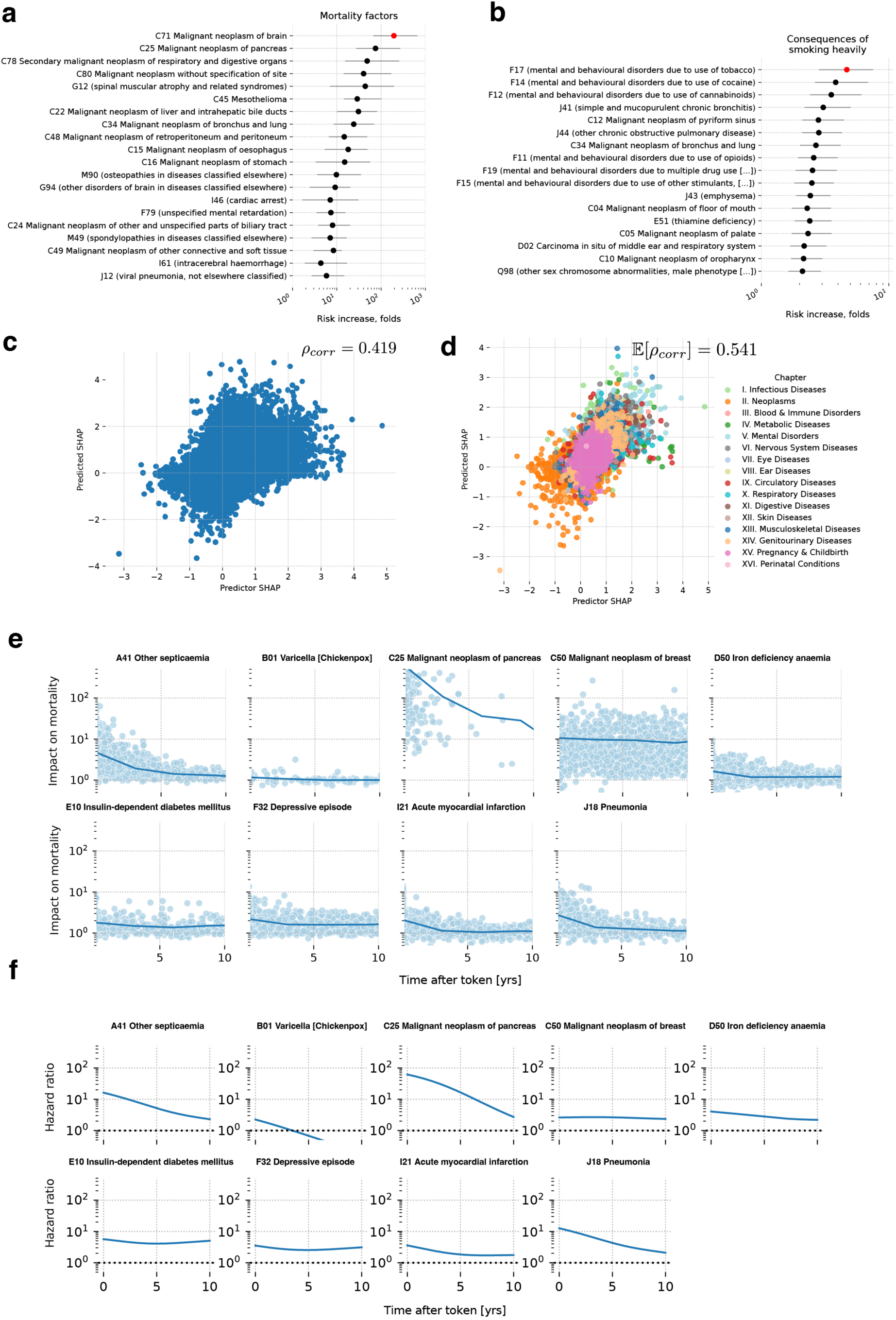
Analysis of estimated influences among diagnoses. **a**, Strongest SHAP predictors of increased mortality risk. Points show the median effect, lines extend to the 25 and 75 percentiles. **b,** Tokens with the largest risk increase, attributed to thes “Smoking - high” lifestyle token. **c,** Scatter plot of pairs “SHAP value (logarithmic fold change) of token X predicting diagnosis Y” and vice versa for all possible pairs of tokens. The value on top denotes the Pearson correlation. **d,** Same as **c**, but restricted to pairs within the same ICD-10 chapters (indicated by colour). **e,** SHAP values (fold change) for mortality (y-axis) as a function of time after diagnosis of 9 different diseases (x-axis). **f,** Nelson-Aalen analysis of the hazard rates for the same diseases shown in **e** confirming trend and magnitude of SHAP effects.

**Extended Data Figure 14.**
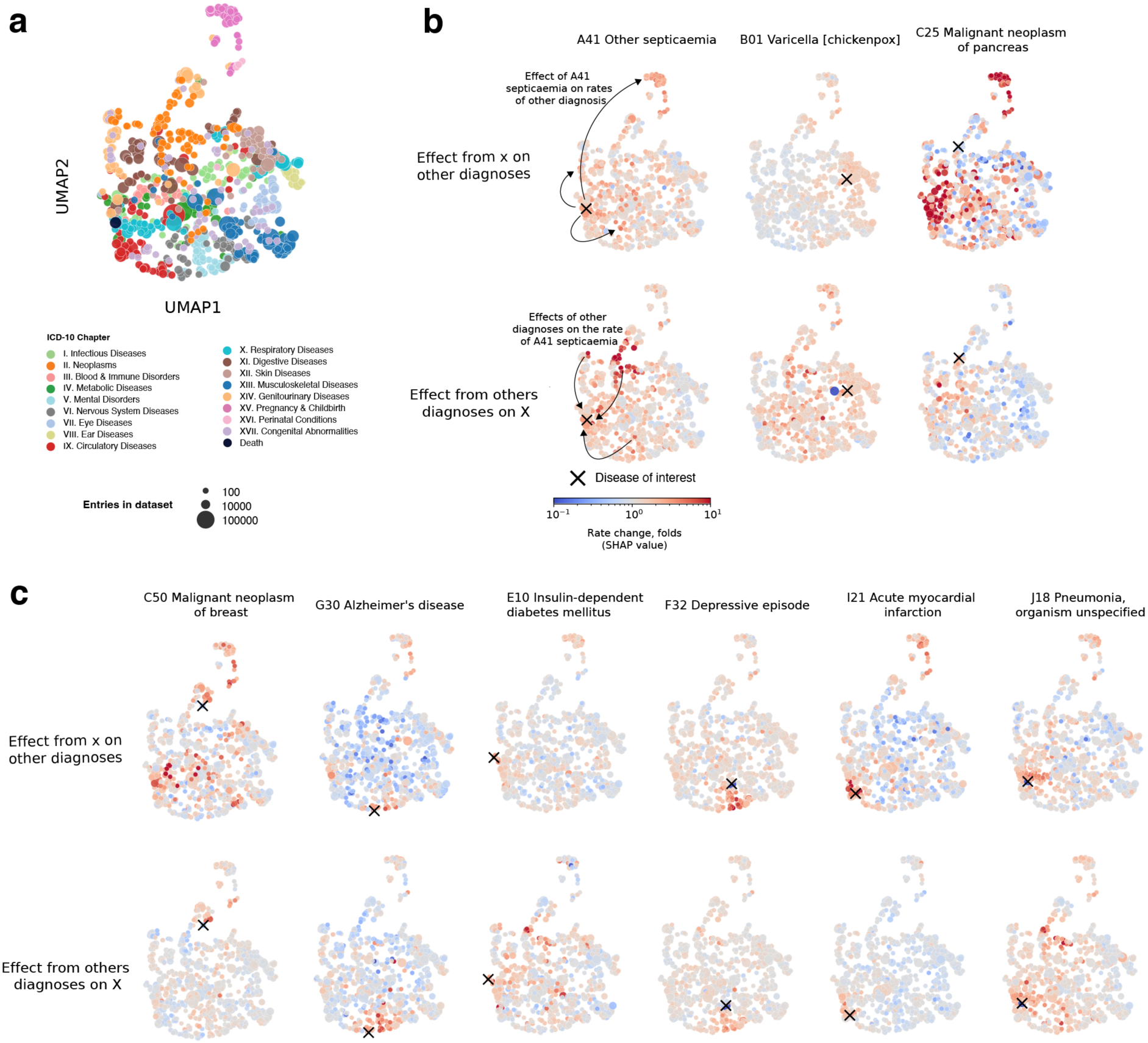
Relation of token embedding space and SHAP effects. **a.** Disease embedding UMAP, coloured by the disease ICD-10 chapter. **b.** UMAP scatter plot, coloured by the SHAP disease rate change for the disease of interest, denoted by a cross marker. According to the SHAP analysis, diseases with similar embeddings tend to have more effect on the predicted rate of each other. Top row, the effect of the selected disease on the rate of other subsequent diseases. Bottom row, the effect of other diseases on the selected disease. **c.** Same as **b**, more diseases.

**Extended Data Figure 15.**
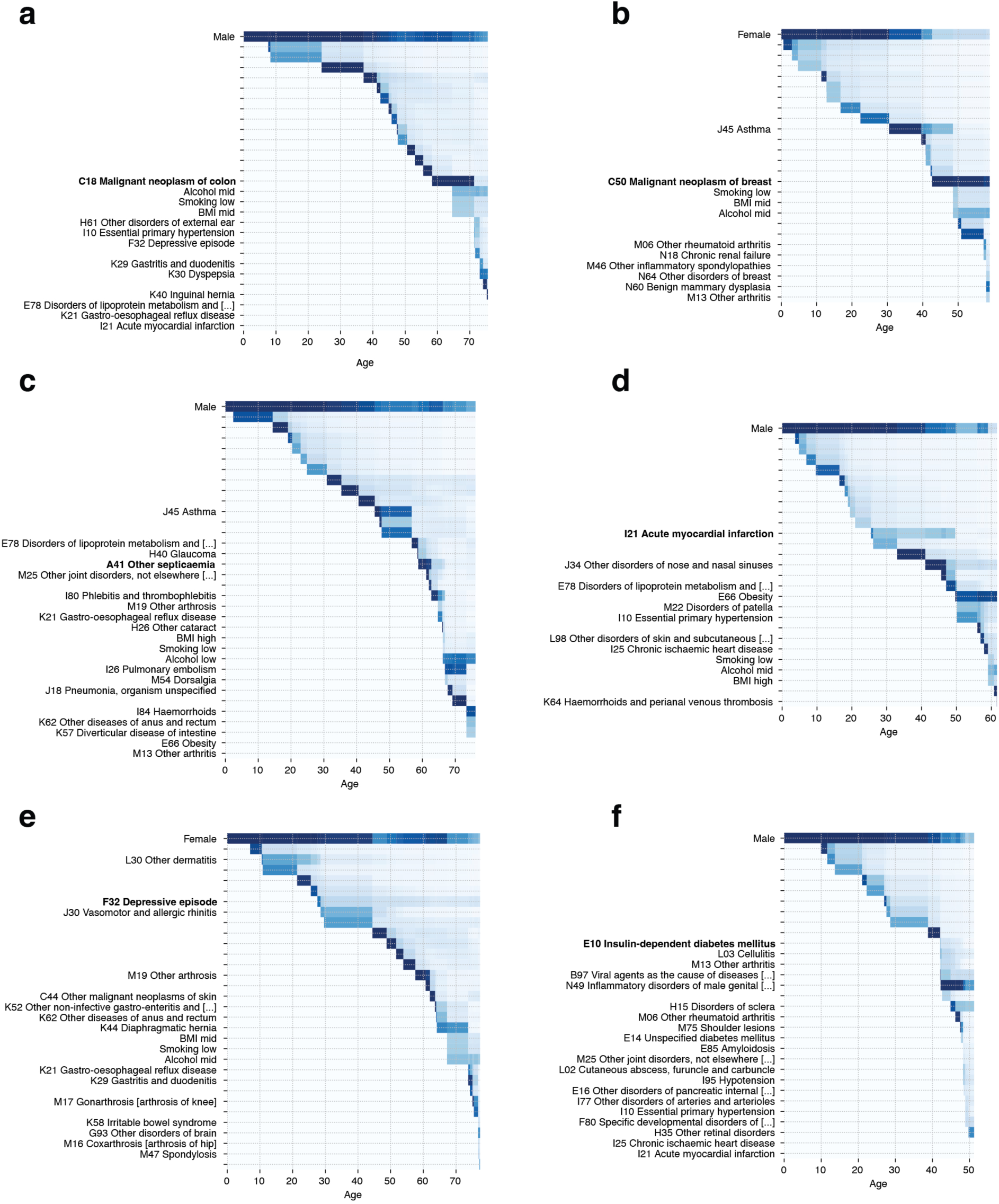
Attention maps indicate temporal effects of prior diagnoses. **a-f**. Attention weights for 6 selected trajectories representative of the token highlighted on the y-axis. Shown is the maximum attention across the 12 layers and 12 heads utilised by Delphi-2M. Darker values indicate higher attention (range 0-1). Typically the last (token, age) pair is most strongly attended to, after which attention drops. Sex and selected other inputs are attended to for more extended periods. Rows without labels are “No event” tokens.

**Extended Data Figure 16.**
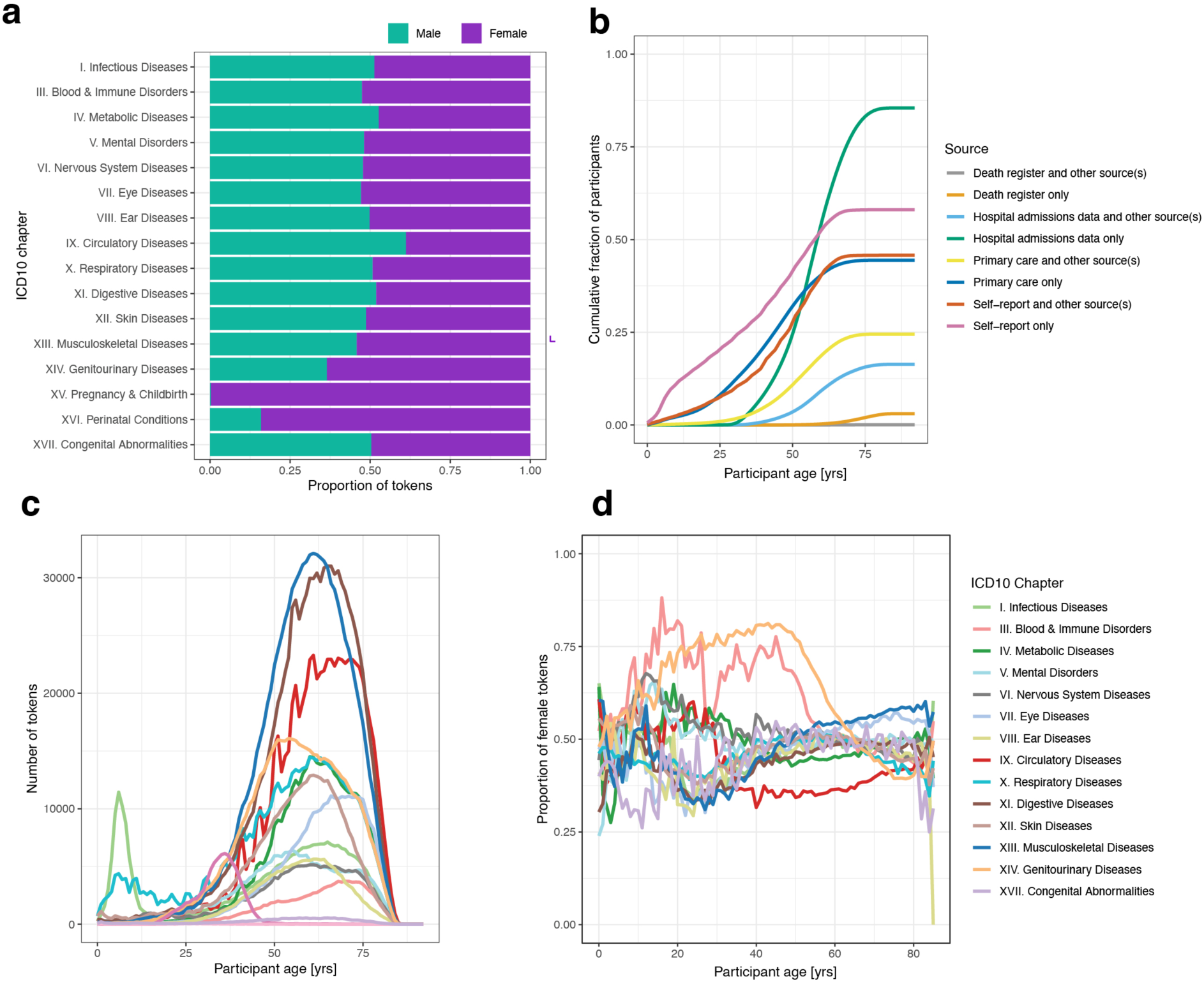
ICD10 disease token biases. **a,** The proportion of the total disease token set scaled by the number of samples, comparing female and male samples across the ICD-10 chapters. **b,** Participant age against the cumulative sum of the fraction of participants with disease tokens across 8 primary data sources. **c,** Participant age against the total number of disease tokens across ICD-10 chapters. **d,** Participant age against the proportion of tokens in females across the ICD-10 chapters.

**Extended Data Figure 17.**
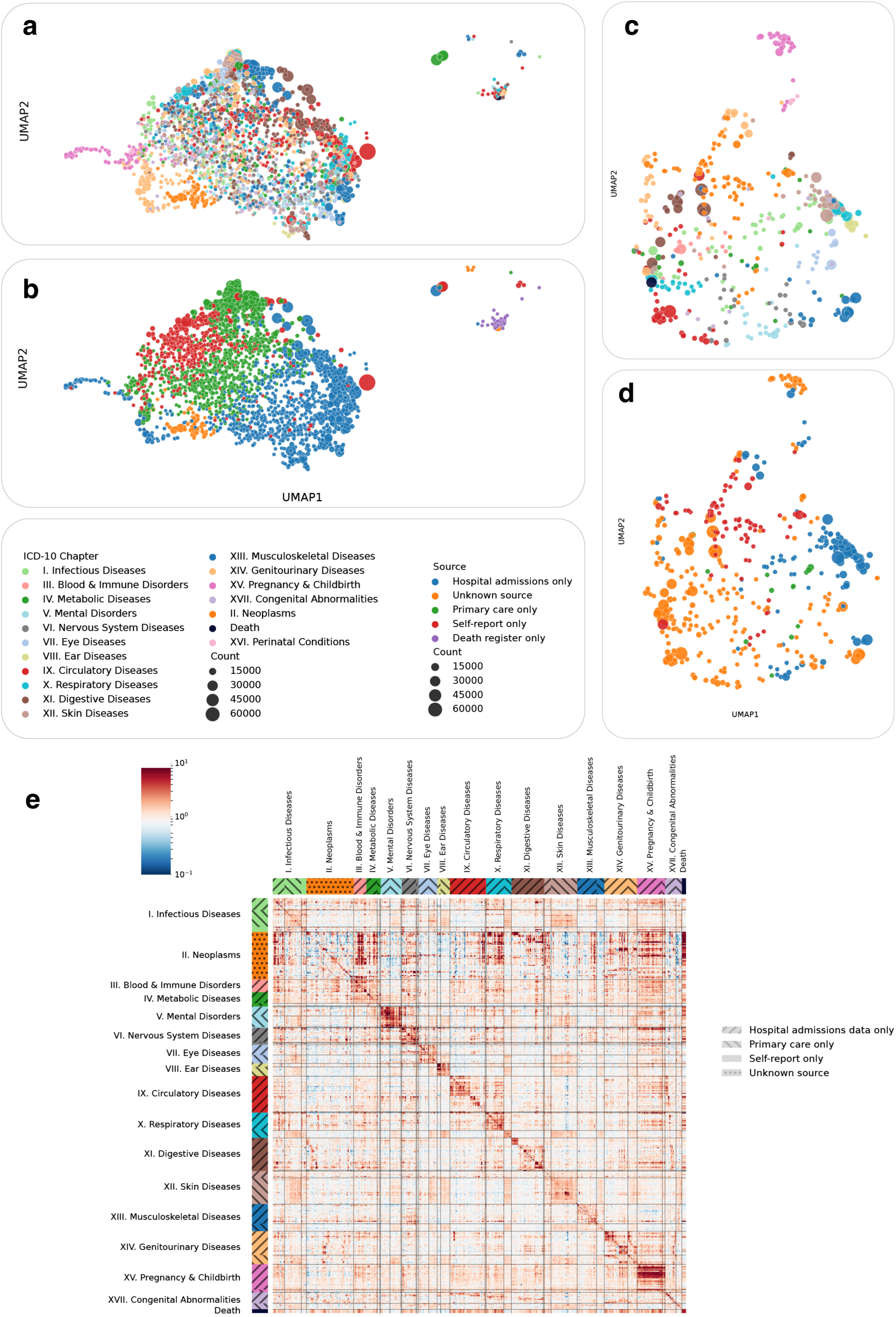
Token source-related biases. Non-random missingness may cause biases in predictions even when courses are not explicitly provided to the model. **a.** Disease embedding UMAP for a Delphi model with explicit token sources (e.g. “Common cold (self-reported)” and “Common cold (hospital records)” are separate tokens), tokens coloured by ICD-10 chapters. **b.** Same as **a**, coloured by token source. **c.** Same as **a**, but for the standard Delphi-2M model. Only tokens with more than 75% of all entries from one source are shown. **d**. Same as **c**, coloured by primary token source. **e**. SHAP value matrix (similar to Figure 4c), with tokens grouped by chapter and primary source.

**Extended Data Figure 18.**
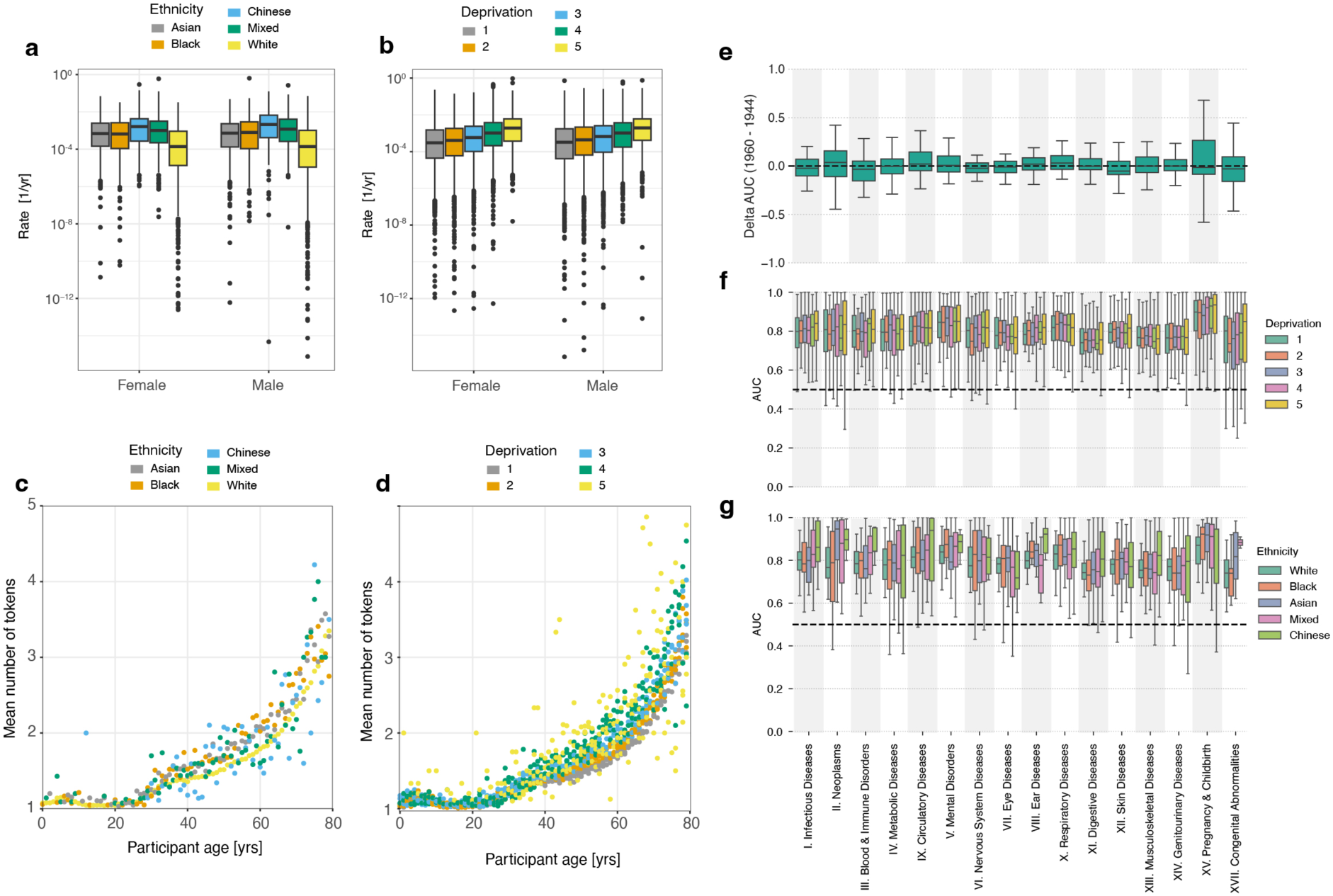
Effects of ethnicity and deprivation. **a,** Modelled rate per year separated by sex and ethnic background. **b,** Modelled rate per year separated by sex and Townsend deprivation index bins (increasing for greater deprivation index values). The boxplots in **a** and **b** feature median as the center line, the box from the first to the third quartile, the whiskers for 1.5x IQR and the outliers. **c-d,** Average number of disease tokens per year, shown for different ethnicities (**с**) and deprivation indices (**d**). **e.** Average validation AUC across 5-year age groups ranging from 40 to 80 years of age, aggregated by the corresponding ICD chapters. Difference between average AUCs calculated for participants with birth years before 1944 and after 1960. **b.** Average per-chapter AUC, participants stratified by deprivation index (**f**) and ethnicity (**g**).

**Extended Data Figure 19.**
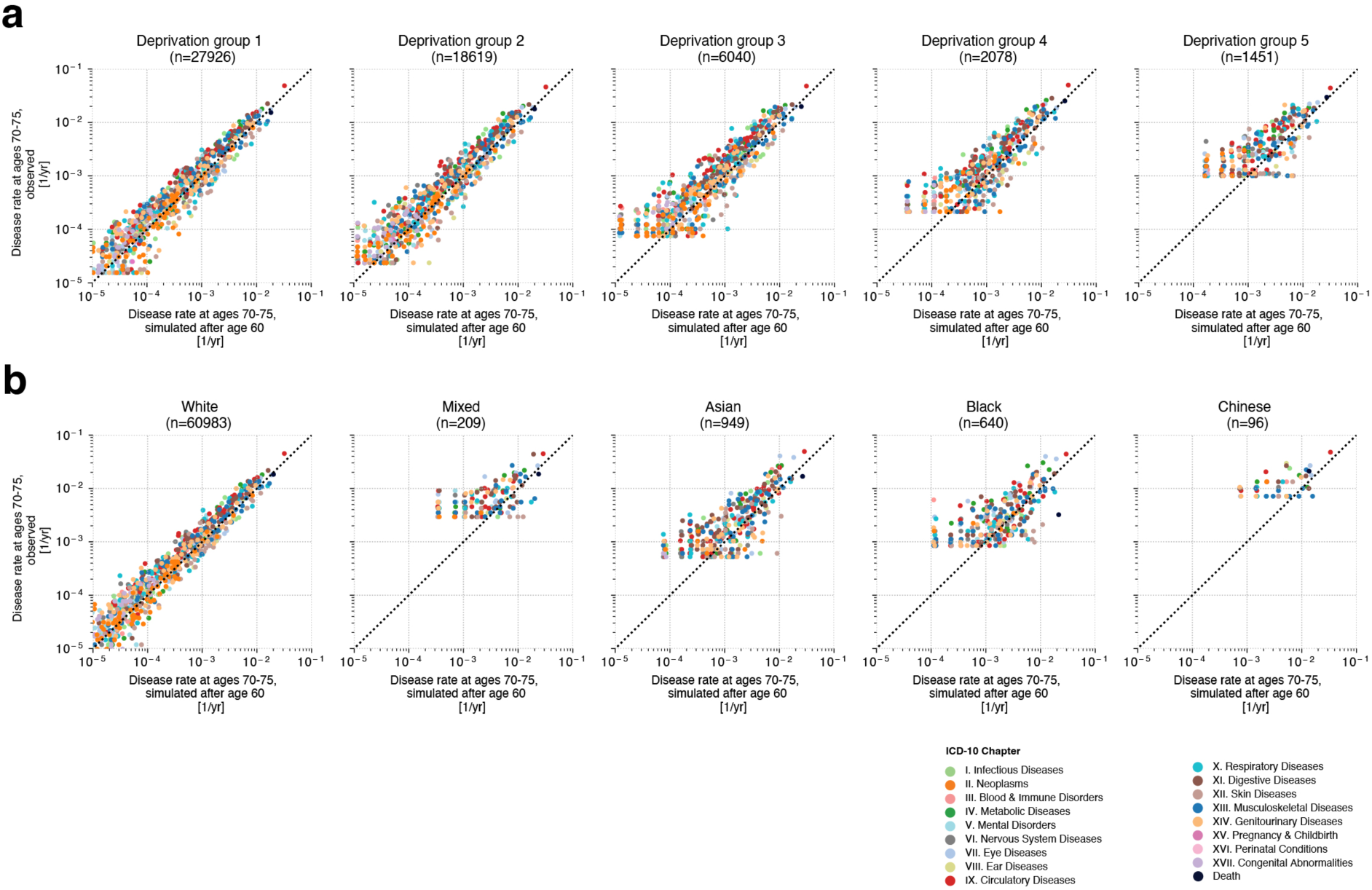
Stratification analysis for generated trajectories. Simulated and observed disease rates recorded between ages 70-75 in UK Biobank validation data. Simulations use data until the age of 60. **a,** Scatter plots of simulated and observed rates in participant groups split by Townsend deprivation index. **b,** Scatter plots of simulated and observed disease rates stratified by self-reported ethnicity. Each dot is an ICD-10 diagnosis coloured by the disease chapter.

## References

1. Nyberg, S. T. et al. Association of healthy lifestyle with years lived without major chronic diseases. JAMA Intern. Med. 180, 760–768 (2020).

2. Link, B. G. & Phelan, J. Social conditions as fundamental causes of disease. J. Health Soc. Behav. **Spec** **No**, 80–94 (1995).

3. Zhu, Z. et al. Causal associations between risk factors and common diseases inferred from GWAS summary data. Nat. Commun. 9, 224 (2018).

4. Mukama, T. et al. Risk-Adapted Starting Age of Screening for Relatives of Patients With Breast Cancer. JAMA Oncol 6, 68–74 (2020).

5. Tian, Y. et al. Calculating the Starting Age for Screening in Relatives of Patients With Colorectal Cancer Based on Data From Large Nationwide Data Sets. Gastroenterology 159, 159–168.e3 (2020).

6. Quan, H. et al. Updating and validating the Charlson comorbidity index and score for risk adjustment in hospital discharge abstracts using data from 6 countries. Am. J. Epidemiol. 173, 676–682 (2011).

7. D’Agostino, R. B., Sr et al. General cardiovascular risk profile for use in primary care: the Framingham Heart Study: The Framingham heart study. Circulation 117, 743–753 (2008).

8. Hippisley-Cox, J. & Coupland, C. Development and validation of risk prediction algorithms to estimate future risk of common cancers in men and women: prospective cohort study. BMJ Open 5, e007825 (2015).

9. Bray, F. et al. Global cancer statistics 2022: GLOBOCAN estimates of incidence and mortality worldwide for 36 cancers in 185 countries. CA Cancer J. Clin. (2024) doi:10.3322/caac.21834.

10. Watt, T. et al. Health in 2040: projected patterns of illness in England. The Health Foundation (2023).

11. Jung, A. W. & Gerstung, M. Bayesian Cox regression for large-scale inference with applications to electronic health records. aoas 17, 1064–1085 (2023).

12. Vaswani, A., et al. Attention Is All You Need. arXiv [cs.CL] Preprint at http://arxiv.org/abs/1706.03762 (2017).

13. Brown, T. et al. Language models are few-shot learners. Adv. Neural Inf. Process. Syst. 33, 1877–1901 (2020).

14. Gemini Team et al. Gemini: A Family of Highly Capable Multimodal Models. arXiv [cs.CL] (2023).

15. Touvron, H. et al. LLaMA: Open and Efficient Foundation Language Models. arXiv [cs.CL] (2023).

16. Ouyang, L. et al. Training language models to follow instructions with human feedback. arXiv [cs.CL] (2022).

17. OpenAI et al. GPT-4 Technical Report. arXiv [cs.CL] (2023).

18. Rasmy, L., Xiang, Y., Xie, Z., Tao, C. & Zhi, D. Med-BERT: pretrained contextualized embeddings on large-scale structured electronic health records for disease prediction. NPJ Digit Med 4, 86 (2021).

19. Li, Y. et al. BEHRT: Transformer for electronic health records. Sci. Rep. 10, 7155 (2020).

20. Li, Y. et al. Hi-BEHRT: Hierarchical Transformer-based model for accurate prediction of clinical events using multimodal longitudinal electronic health records. IEEE J. Biomed. Health Inform. 27, 1106–1117 (2023).

21. Yang, Z., Mitra, A., Liu, W., Berlowitz, D. & Yu, H. TransformEHR: transformer-based encoder-decoder generative model to enhance prediction of disease outcomes using electronic health records. Nat. Commun. 14, 7857 (2023).

22. Placido, D. et al. A deep learning algorithm to predict risk of pancreatic cancer from disease trajectories. Nat. Med. 29, 1113–1122 (2023).

23. Savcisens, G. et al. Using sequences of life-events to predict human lives. Nat. Comput. Sci. 1–14 (2023).

24. Kraljevic, Z. et al. Foresight—a generative pretrained transformer for modelling of patient timelines using electronic health records: a retrospective modelling study. *Lancet Digit*. Health 6, e281–e290 (2024).

25. Radford, A. et al. Language Models are Unsupervised Multitask Learners. Preprint at https://d4mucfpksywv.cloudfront.net/better-language-models/language-models.pdf.

26. Hoffmann, J. et al. Training Compute-Optimal Large Language Models. arXiv [cs.CL] (2022).

27. Garg, M. et al. Disease prediction with multi-omics and biomarkers empowers case-control genetic discoveries in the UK Biobank. Nat. Genet. 56, 1821–1831 (2024).

28. Lundberg, S. M. & Lee, S.-I. A Unified Approach to Interpreting Model Predictions. in Advances in Neural Information Processing Systems (eds. Guyon, I. et al.) vol. 30 (Curran Associates, Inc., 2017).

29. Fry, A. et al. Comparison of Sociodemographic and Health-Related Characteristics of UK Biobank Participants With Those of the General Population. Am. J. Epidemiol. 186, 1026–1034 (2017).

30. Shuster, K., Poff, S., Chen, M., Kiela, D. & Weston, J. Retrieval Augmentation Reduces Hallucination in Conversation. arXiv [cs.CL] (2021).

31. Imrie, F., Rauba, P. & van der Schaar, M. Redefining digital health interfaces with large Language Models. arXiv [cs.CL] (2023).

32. Sudlow, C. et al. UK biobank: an open access resource for identifying the causes of a wide range of complex diseases of middle and old age. PLoS Med. 12, e1001779 (2015).

33. Schmidt, M. et al. The Danish National Patient Registry: a review of content, data quality, and research potential. Clin. Epidemiol. 7, 449–490 (2015).

34. Helweg-Larsen, K. The Danish Register of Causes of Death. Scand. J. Public Health 39, 26–29 (2011).

35. Pedersen, M. K. et al. A unidirectional mapping of ICD-8 to ICD-10 codes, for harmonized longitudinal analysis of diseases. Eur. J. Epidemiol. 38, 1043–1052 (2023).

36. Aalen, O. Nonparametric Inference for a Family of Counting Processes. Ann. Stat. 6, 701–726 (1978).

37. Nelson, W. Theory and applications of hazard plotting for censored failure data. Technometrics 14, 945 (1972).

38. Hippisley-Cox, J., Coupland, C. & Brindle, P. Development and validation of QRISK3 risk prediction algorithms to estimate future risk of cardiovascular disease: prospective cohort study. BMJ 357, j2099 (2017).

39. Imrie, F., Cebere, B., McKinney, E. F. & van der Schaar, M. AutoPrognosis 2.0: Democratizing diagnostic and prognostic modeling in healthcare with automated machine learning. *arXiv [cs.LG]* (2022) doi:10.1371/journal.pdig.0000276.

40. Anatürk, M. et al. Development and validation of a dementia risk score in the UK Biobank and Whitehall II cohorts. BMJ Ment Health 26, (2023).

41. McInnes, L., Healy, J. & Melville, J. UMAP: Uniform Manifold Approximation and Projection for Dimension Reduction. arXiv [stat.ML] (2018).

